# A prospective cohort study of unselected nulliparous women with a nested randomized controlled trial of screening using the sFLT1:PlGF ratio, ultrasound and maternal characteristics and intervention using enhanced monitoring and early delivery: study protocol for the POPS2 cohort and randomized controlled trial

**DOI:** 10.64898/2026.04.29.26352041

**Authors:** Gordon C. S. Smith, Amy Sutton-Cole, Ellen Dyer, Ulla Sovio, Emma Cook, Ian R White, D. Stephen Charnock-Jones

## Abstract

**Introduction:** Current UK guidelines recommend measurement of symphyseal fundal height and measurement of maternal blood pressure and urinalysis, with the aim of detecting women at increased risk of fetal growth restriction (FGR) and preeclampsia. Between 2008 and 2013 we conducted a prospective cohort study recruiting 4,512 nulliparous women at the Rosie Hospital, Cambridge, where we performed serial ultrasonic imaging and serial blood sampling and generated a novel screening test for preeclampsia and FGR. The method involved measuring the ratio of two placental biomarkers (soluble fms-like tyrosine kinase receptor-1 [sFLT1] and placenta growth factor [PlGF]) at ∼36 weeks of gestational age (wkGA) and combining the result with maternal characteristics and ultrasonic imaging. Women who screened positive had a ∼50% risk of a composite outcome, consisting of preeclampsia ± delivery of a baby with a birth weight <3rd percentile for sex and gestational age ± perinatal morbidity or death. It is plausible that screening and intervention using this method might improve pregnancy outcome.

**Methods and analysis:** Nulliparous women with an apparently normal singleton pregnancy will be recruited at their dating ultrasound scan. Blood will be obtained at this visit, at their anomaly scan (20wkGA), and at two research appointments (28wkGA and 36wkGA) when research ultrasound scans will be performed. Blood for DNA will be obtained from the father of the baby where possible and the placenta will be sampled following birth. At 36wkGA, women will be consented for participation in the randomised controlled trial (RCT) element of the study and their risk of term preeclampsia and FGR will be assessed using the novel approach. Women who screen high-risk will then be randomly allocated to either having the result revealed or masked. Women randomised to having the result revealed will be offered early delivery and/or enhanced monitoring. Where the result is masked, there will be no communication between the research team and the participant, and she will continue to receive routine care at the Rosie Hospital. The primary outcome is a composite of preeclampsia, FGR and perinatal morbidity and mortality. The study will also generate data and biological samples to support future research in novel screening methods and disease mechanisms.

**Ethics and dissemination:** The study received ethical approval from the East of England Research Ethics Committee. All women provide written informed consent to participate in the cohort. Women provide a second written informed consent to participate in the RCT. The study results will be disseminated by presentation at international conferences and publication in peer reviewed journals.

**Trial registration:** 07/10/2019 ISRCTN12181427 (https://doi.org/10.1186/ISRCTN12181427)

**Strengths and limitations of the study:** 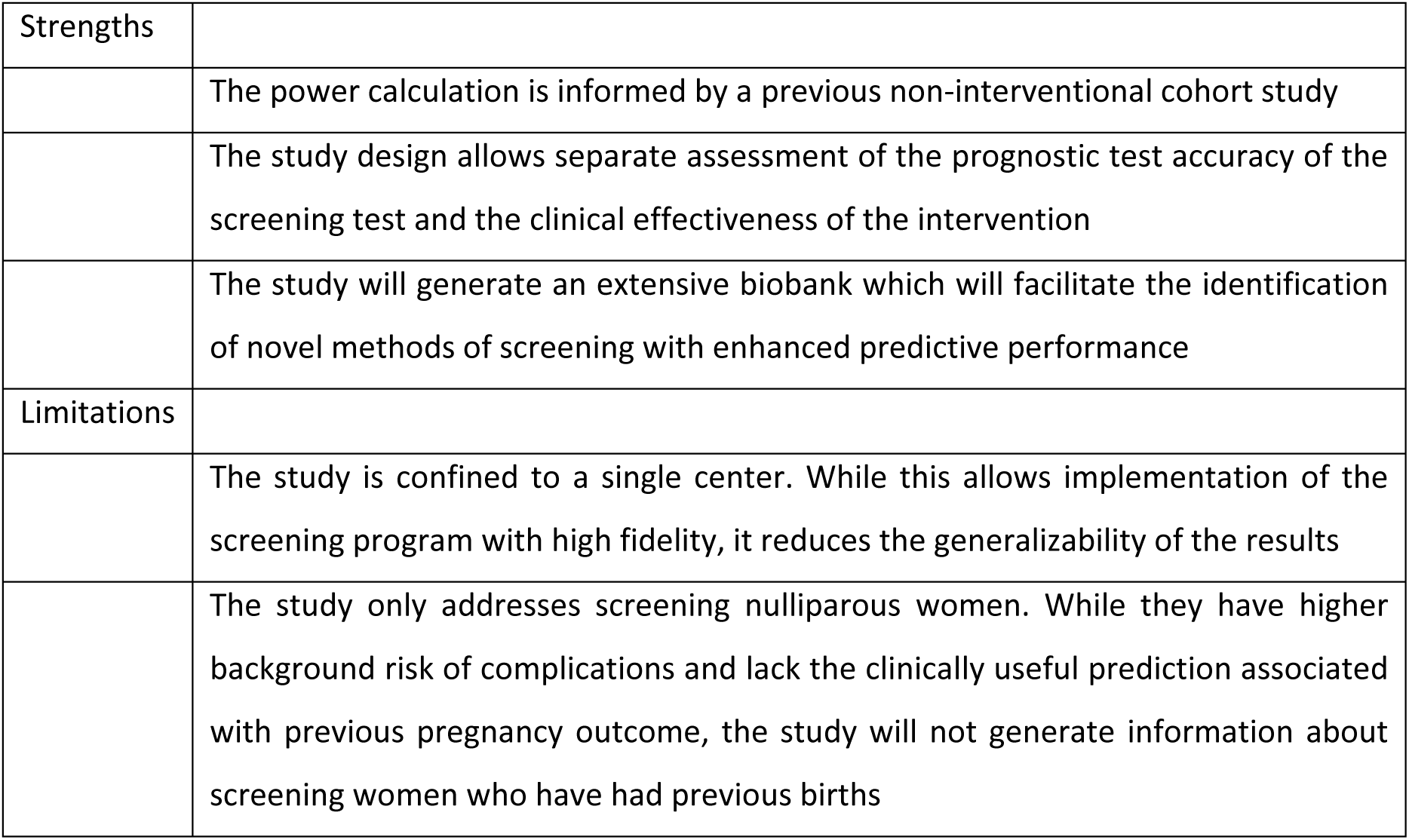

## Background and rationale

Maternal and perinatal deaths account for 6-7% of all deaths globally, approximately equivalent to half of the global burden of deaths caused by cancer(1). Placental dysfunction is implicated in the aetiology of many of the associated conditions, including preeclampsia, fetal growth restriction (FGR), spontaneous preterm birth and stillbirth(2). The approaches to screening for these outcomes include methods which are relatively crude and ineffective. For example, the UK’s National Institute for Health and Care Excellence (NICE) recommended method of screening for FGR is to measure the external size of the uterus with a tape measure, despite the fact that surveillance based on this approach has a sensitivity to detect small infants of ∼20%(3). An alternative is to perform late pregnancy ultrasonic fetal biometry in all women. However, a meta-analysis of eight RCTs of universal ultrasound recruiting ∼35,000 women showed no evidence of benefit(4). Implementation of universal late pregnancy ultrasound in France was associated with dramatic increases in morbidity in false positives, principally through iatrogenic prematurity(5).

In a 2012 review in PLoS Medicine(6), we outlined key methodological weaknesses in the literature:

- screening tools were evaluated in interventional trials in the absence of high quality information on their diagnostic effectiveness
- screening tests were not coupled to an effective intervention
- trials were underpowered

The review proposed a plan to generate novel clinically effective methods of screening:

- trials should be preceded by high quality studies of diagnostic effectiveness
- initial efforts should focus on screening for complications at term
- the most efficient trial design is to screen all women and randomise high-risk women to intervention or routine care
- initial efforts should focus on nulliparous women(7), as they have a higher a *priori* risk of adverse pregnancy outcome. They lack information on complications during previous pregnancies(8), one of the most discriminating predictors of preeclampsia and FGR.

In 2007, we designed the Pregnancy Outcome Prediction study (POPS), a prospective cohort study of nulliparous pregnant women in the Rosie Hospital, Cambridge. The protocol has been published(7), the first output of the study described the conduct of the cohort(3), and a review article describes the biological samples (9). The study followed the first part of the plan outlined in the PLoS Medicine review and has generated a screening test for complications at term. Combining the data from two previous publications arising from the POPS cohort(10, 11), we have created a screening test (described in detail below) which is conducted at around 36 weeks of gestational age (wkGA) and has a high positive predictive value, but has a sensitivity of less than 50%. This means that, while screening and intervention may be beneficial for individual women, better tests are required to yield substantial reductions in the population incidence. Moreover, POPS2 is confined to screening and intervention for preeclampsia and FGR near term and does not address the problem of preeclampsia and FGR associated with preterm birth. Women who are identified as high-risk at 28wkGA have an absolute risk of preterm delivery of a small infant of about 20%(11). Hence, for one true positive there would be four false positives. Consequently, use of this test at 28wkGA has the potential to cause net harm through iatrogenic prematurity in the false positives. This background information informs the objectives of the study.

### Objectives

We aim to perform a nested RCT within a prospective cohort study of nulliparous women with a singleton pregnancy to determine (a) whether screening and intervention using a novel method for assessing the risk of preeclampsia and fetal growth restriction at term correctly identifies women at increased risk of complications, and (b) whether intervention in high-risk women improved the outcome for the mother and/or infant. Our secondary aim is to use the data and biological samples from the cohort study to generate better performing screening tests for preeclampsia, FGR and other adverse outcomes of pregnancy. The secondary aim will address the two weaknesses described above, namely, the less than 50% sensitivity of our current test to detect preeclampsia at term and the absence of screening tools with a high positive predictive value for preeclampsia and FGR resulting in preterm birth. However, we believe that the current screening test has the potential to prevent adverse outcomes using technology that is currently available and licensed for clinical use. Given that it could take many years to develop, validate and obtain regulatory approval for novel tests, we believe an RCT of our existing method is justified now. A further goal of the extensive collection of biological samples is to use these to better understand the associations and mechanisms linking pregnancy to health and disease in the mother and offspring.

### Trial design

The study is a prospective cohort study with a nested, parallel group RCT where women screening high-risk will be allocated in a ratio of 1 to 1 to intervention or routine care, to test the superiority of intervention among women who screen high-risk.

## METHODS: PARTICIPANTS, INTERVENTIONS AND OUTCOMES

### Study setting

The study will take place at a single centre, The Rosie Hospital, Cambridge, UK, which is an academic maternity hospital providing care for about 5000 to 6000 deliveries per annum, serving the local population of Cambridge as well as providing tertiary level obstetric, maternal-fetal medicine and neonatal care for hospitals in the East of England, UK.

### Eligibility criteria

#### Cohort study

Eligible women will be those who are nulliparous (no previous births >23wkGA) with a single viable fetus at the dating ultrasound scan with an ultrasonic estimated gestational age <18wkGA. Exclusion criteria for the cohort study will be those who are unable to provide consent, maternal age <16, previous births >23wkGA, multiple pregnancy, and a non-viable fetus or a fetus with a major anomaly or nuchal translucency >3.5mm at the time of the dating ultrasound.

#### Randomized controlled trial

The RCT is nested within the cohort, i.e. all women participating in the RCT will be cohort members but not all cohort members will participate in the RCT. Inclusion criteria for the RCT are women who were recruited to the cohort study and who attended for their 36wkGA visit. Exclusion criteria are (a) participants where the 36wkGA research scan demonstrated one of the findings which mandates that the result would be revealed (see below), (b) any participant not planning to deliver at the Rosie Hospital, or (c) any participant who had already been offered a date for medically indicated delivery within seven days of their 36wkGA visit.

### Who will recruit participants and document their consent?

#### Cohort

In the Rosie Hospital, women who require a dating ultrasound scan will be identified either by receipt of a “Notification of Pregnancy” form or when a woman has confirmation of pregnancy in primary care and contacts the hospital to arrange a dating scan. The patient information leaflet will be sent with the appointment details.

The dating scan will be performed as routine. Any adverse findings on scan, e.g. delayed miscarriage or suspected fetal abnormality, will follow the normal referral pathway. The sonographer conducting the scan will confirm that the woman has not had a previous birth beyond 23wkGA and that she has a viable singleton pregnancy <18wkGA at the time of the scan. The sonographer will ask the woman if she is happy to meet with the member of the research team to discuss the study. If they agree, the sonographer will inform the research staff that the woman is potentially interested in participating.

Women will also be alerted to the study through other means. This will cover the situation where the timing of the research scan lists was inconvenient. For example, we will place posters in prominent areas of the Rosie Hospital, including the ultrasound department, we will put a notice on the Rosie Hospital website, and we will also use social media sites (such as Facebook and X, (previously Twitter) to disseminate this information. These notices will alert women who were not approached for recruitment at the time of their dating ultrasound and provide the contact details for them to get in touch and arrange an appointment to come and discuss the study. Moreover, the patient information leaflet will be sent to all eligible women and it will detail how they can participate in the study if they are not approached at the time of the dating ultrasound.

Consent will be given following discussion of the study with an appropriately trained member of staff, such as a research midwife, research nurse, or research sonographer.

The following are some of the key points to be raised during the consent process:

- Participation in the study is voluntary, consent can be withdrawn at any time during the study, and withdrawal will not affect their routine care
- The purpose of the study
- The requirements of participating in the study
- That women will not be informed of the results of the 28wkGA and 36wkGA scans except for intra-uterine fetal death, previously undiagnosed and clinically relevant congenital anomaly, severely reduced amniotic fluid level, placenta praevia, grossly abnormal umbilical blood flow, and non-cephalic presentation (36wkGA)
- All information will be treated as strictly confidential
- These analyses could include genetic studies and studies done by commercial entities
- Participation in the study will not affect their routine care, except that they will be referred for assessment by the medical team if either of the scans demonstrated one of the findings described above
- That they will be asked at the time of their 36wkGA visit whether they want to participate in the RCT. This will involve giving them a patient information leaflet at their 28wkGA visit, discussing the information on it, and signing a consent from at their 36wkGA visit. The conduct of the RCT is described below.

Information relating to data handling and biological samples is detailed below.

The signed consent form has three copies, one will be retained for the study records, one placed in the patient’s hand-held notes, and the third scanned for recording in the hospital’s Electronic Medical Record (EMR, specifically the EPIC system). A letter will be sent to the woman’s GP informing them of participation in the study.

When women give consent, they will be directed to the NIHR Cambridge Clinical Research Facility for blood samples to be obtained (see below) and to make their next appointment.

#### Father

When the father attends for one of the research ultrasounds, consent will be sourght to take blood by an appropriately trained member of staff, such as a research midwife or research sonographer.

The following are some of the key points to be raised during the consent process:

- Participation in the study is voluntary, consent can be withdrawn at any time during the study, and withdrawal will not affect their routine care
- The purpose of the study
- The requirements of participating in the study
- All information will be treated as strictly confidential
- These analyses could include genetic studies and studies done by commercial entities
- We will not report any result that allows identification of your baby’s paternity

Information relating to data handling and biological samples is detailed below.

#### Randomized controlled trial

Eligible women will be consented by an appropriately trained and qualified member of the research team, for example, a sonographer or a research midwife. If for any reason recruitment to the RCT element of the study is not possible (for example, in the unlikely event of failure of the machine which performs the sFLT1 and PlGF assays), women will still have their 36wkGA scan and blood sample for the cohort element of the study. For women who are approached to consent to the RCT, the discussion will include the following information:

- The blood sample will be analysed using a test which is believed to indicate the health of the placenta
- The blood test is in use in other clinical situations but it is not currently recommended as a screening test, i.e. it is currently only used in situations where a woman has a problem
- We have evidence that the test, when combined with the scan results and the mother’s own past history, identifies women at an increased risk of complications
- The aim of the study is to determine whether performing the test and acting on the results leads to a lower rate of complications
- Consenting to the study means that we will perform the test on her blood sample
- If she is deemed high-risk on the basis of the test, she will then be randomised to routine care or to being contacted to discuss how to manage the increased risk of complications
- The main intervention will be to offer earlier delivery of the baby by induction of labour
- This will be offered from 37+0, and will be strongly recommended no later than 39+0. Delivery at 37wkGA will be strongly recommended if the blood test and/or scan were very strongly suggestive of a problem
- If she screens positive and is randomised to intervention, she will be contacted within 7 days or fewer
- About 3 to 4 women per 100 will be contacted
- She should not assume that she is low risk if she is not contacted, as some women (about 3 to 4 women per 100) will have screened high-risk and been randomised to routine care
- Hence, if she has any concerns about any aspect of the pregnancy (for example her baby’s movements, bleeding, symptoms), she should contact her midwife or doctor as she normally would.
- Not being contacted by the research team does not mean that she can assume that she is not at risk of complications.

The signed consent form has three copies, one will be retained for the study records, one placed in the patient’s hand-held notes, and the third study copy will be scanned for recording in EPIC. A letter will be sent to the woman’s GP informing them of participation in the RCT element of the study.

### Additional consent provisions for collection and use of participant data and biological specimens

#### Cohort study

The following are key points to be raised during the consent process for recruitment to the cohort:

- Participation will involve research staff accessing medical notes and linking electronic clinical records to their research data, including longer term outcomes (such as subsequent pregnancies [mother] and educational outcome [child])
- Their personal identifiable information will be held securely. Their clinical and research data will only be shared with other researchers if we are certain that it is anonymous and that they cannot be identified.
- They are providing broad consent for the analysis of their blood and other samples, to be used in any way that enhances our understanding of normal and complicated pregnancy and how this influences health and wellbeing in later life.
- These analyses could include genetic studies and studies done by commercial entities.

### Interventions

Eligible women who consent to the RCT element of the study will have the sFLT1:PlGF ratio (the concentration of soluble fms-like tyrosine kinase receptor 1 divided by the concentration of placenta growth factor, measured on a Roche Cobas assay platform(10)) measured in their 36wkGA serum sample. The risk status will be based on the combination of the exact level of the ratio plus other maternal characteristics and ultrasound scan findings.

Screen positive will be defined as ≥1 of the following:

1. sFLT1:PlGF ratio >110
2. sFLT1:PlGF ratio >38 + 36wkGA ultrasonic estimated fetal weight (EFW) <10th percentile(11)
3. sFLT1:PlGF ratio >38 + 20wkGA uterine artery Doppler pulsatility index >90th percentile(10)
4. sFLT1:PlGF ratio >38 + increased risk of preeclampsia using an adapted version of the NICE Guideline Antenatal Care defined as ≥1 of the following:

a. chronic kidney disease
b. autoimmune disease, such as systemic lupus erythematosus or antiphospholipid syndrome
c. type 1 or type 2 diabetes
d. chronic hypertension
e. age 40 years or older
f. Body mass index ≥35 kg/m² at first visit

Screen positive women will then be randomly allocated to (a) revealing the result with review by a doctor (usually a consultant in Maternal-Fetal Medicine), or (b) masking the result, with the consequence that the woman receives the same care that she would have received had she not participated in the RCT. Women will be randomised using a computerised system employing stratification on the basis of the main four elements of the risk assessment. The rationale for the choice of the two groups for comparison is that women who have the result masked will have pregnancy care as if they had not had the screening test at all. We will then be able to compare the intervention group with the high-risk masked group to determine whether revealing the result and changing clinical care on the basis of the result reduced the risk of the primary outcome. The study design also allows us to compare outcomes among the women who screened high-risk but were randomised to masking of the result with the women who screened low risk. This comparison will allow assessment of how well the screening approach performs in identifying high-risk women in this population.

### Intervention description

Women who screen positive and are randomised to the intervention arm will be assessed as soon as possible thereafter, with the aim of conducting the assessment within seven days of the blood sample being obtained. The primary intervention is to offer induction of labour with weekly monitoring and to re-offer induction of labour for women who decline. The recommended timing of the induction of labour will depend on which elements of the screening assessment were positive. Women with extreme elevation of the ratio (>110) or those with an elevated ratio (>38) plus a baby that is small for gestational age (SGA) on ultrasound (estimated fetal weight [EFW] <10^th^ percentile) will be recommended to have induction of labour at 37 weeks. All other women will be recommended to have induction of labour at 39 weeks. The doctor assessing the patient will have access to a report containing the results of the screening assessment. For the purposes of the study, the Rosie Hospital has a clinical guideline for the management of screen positive women in the trial and this is available to all staff through the hospital’s intranet.

### Criteria for discontinuing or modifying allocated interventions

The allocated intervention will be either routine care or to reveal the screening test result and to make a recommendation around the timing of induction of labour, and these would apply in all situations. However, a control participant may be recommended to have induction of labour for some other reason, as part of their routine clinical care. Conversely, intervention participants can decline the offer of induction and await the spontaneous onset of labour.

### Strategies to improve adherence to interventions

The medical staff who see high-risk participants randomised to intervention will generally be sub-specialist consultants in Maternal-Fetal Medicine who have had training about the study and are fully acquainted with the details of the clinical guideline. Compliance with the guideline can be assessed at the time of analysis as we predict that the interval from the fourth research visit and birth will be shorter in the intervention group due to the recommendation to induce labour between 37+0 and 39+0 weeks.

### Relevant concomitant care permitted or prohibited during the trial

All other aspects of the care of participants will be as per Rosie Hospital clinical guidelines and no specific care will be mandated or prohibited by participation in the study.

### Provisions for post-trial care

The trial team aims to see each participant who is randomised to intervention in the postnatal period and during the delivery episode, although this may not always be possible. The purpose of the visit will be to check on the patient’s wellbeing and to answer any questions they have in relation to their care. In relation to compensation for harm, Cambridge University Hospitals NHS Foundation Trust, as a member of the NHS Clinical Negligence Scheme for Trusts, accepts full financial liability for harm caused to participants in the study caused through the negligence of its employees and honorary contract holders. The University of Cambridge has an insurance policy in place for negligent harm caused as a result of protocol design and for non-negligent harm arising through participation in the study.

### Outcomes

The study has two primary outcomes and both are composites. The first is one or more of the following: (a) diagnosis of preeclampsia (using the ACOG 2013 definition(12)), (b) perinatal death or perinatal morbidity, or (c) delivery of an infant with a birth weight <3rd percentile for sex and gestational age. Perinatal death is defined as stillbirth or neonatal death. Perinatal morbidity is defined as one or more of the following: a 5 minute Apgar score <7, delivery with metabolic acidosis (defined as a umbilical cord artery or vein pH <7.1 and a base deficit of >10mmol/L) or admission to the neonatal unit (defined as admission <48 hours after birth and discharge ≥48 hours after admission) i.e. either the Special Care Baby Unit (SCBU) or the Neonatal Intensive Care Unit (NICU). The second primary outcome is a subgroup of the first and is defined as one or more of the following: (a) preeclampsia with severe features (ACOG 2013 definition(12)) or (b) perinatal death or severe perinatal morbidity. Severe perinatal morbidity is defined as livebirth associated with hypoxic ischaemic encephalopathy (any grade), use of inotropes, mechanical ventilation, or severe metabolic acidosis (defined as an umbilical cord artery or vein pH <7.0 and a base deficit of >12mmol/L).

Secondary outcomes will include each of the individual elements of the maternal and perinatal composite described above. The following additional secondary outcomes will be studied, based on a previous trial of term induction of labour, the 35/39 trial(13). Maternal: (a) maternal death, (b) maternal stroke, (c) maternal admission to any medical unit (e.g. intensive care unit, stroke unit, coronary care unit, neurosciences critical care unit) (d) placental abruption, (e) cord prolapse, (f) caesarean section, (g) epidural anaesthesia, (h) instrumental vaginal delivery, (i) general anaesthesia, (j) post-partum haemorrhage (any >500ml), (k) post-partum haemorrhage requiring blood transfusion, (l) infection (pyrexia >38°C and treated with a course of antibiotics).

Perinatal: (a) Apgar score at 1 minute, (b) Apgar score at 5 minutes, (c) Apgar score at 10 minutes, (d) umbilical arterial pH and base deficit, (e) umbilical vein pH and base deficit, (f) any/duration of admission to SCBU, (g) any/duration of admission to NICU, (h) fracture (any), (i) seizures (any), (j) prolonged hypotonia (>2h), (k) abnormal level of consciousness (hyperalert, drowsy, lethargic, stupor, decreased response to pain, coma), (l) tube feeding (any duration), (m) head cooling, (n) phototherapy, (o) severe hypoglycaemia (defined as requiring parenteral treatment or drug therapy), (p) birth weight, (q) birth weight percentile for sex and gestational age, (r) stage of hypoxic ischaemic encephalopathy, (s) oxygen administration and/or respiratory support. All diagnoses relate to the delivery admission, i.e. all events until discharge of the mother or baby.

### Participant timeline

Figure 1 summarises the timeline of the cohort and Table 1 summarises the RCT timeline (as per the SPIRIT guideline). Women will be recruited at their dating ultrasound scan, typically performed ∼12wkGA. Where possible blood will be obtained at this visit, at their anomaly scan (20wkGA), and at two research appointments (28wkGA and 36wkGA), when research ultrasound scans will be performed where possible (conducted as per the original POP study(7)). Blood for DNA will be obtained from the father of the baby where available and following informed consent. The scans at 28wkGA and 36wkGA will take place in the NIHR Cambridge Clinical Research Facility. At the 36wkGA visit, eligible women will be approached to ask them to give their consent for participation in the RCT element of the study. Figure 2 illustrates an overview of the design of the RCT element of study.

**Figure 1.**
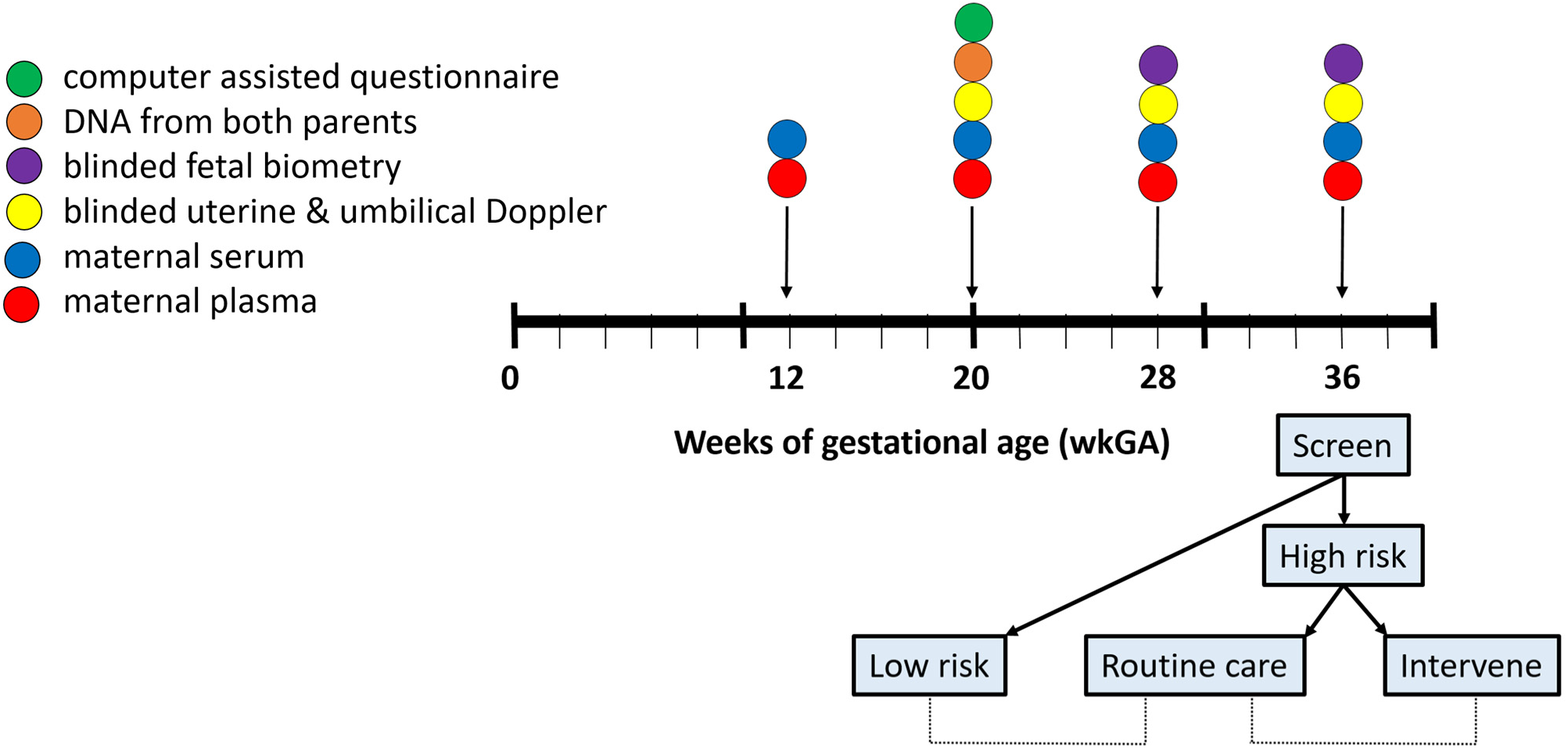
Schematic representation of data and sample collection in the study.

**Figure 2.**
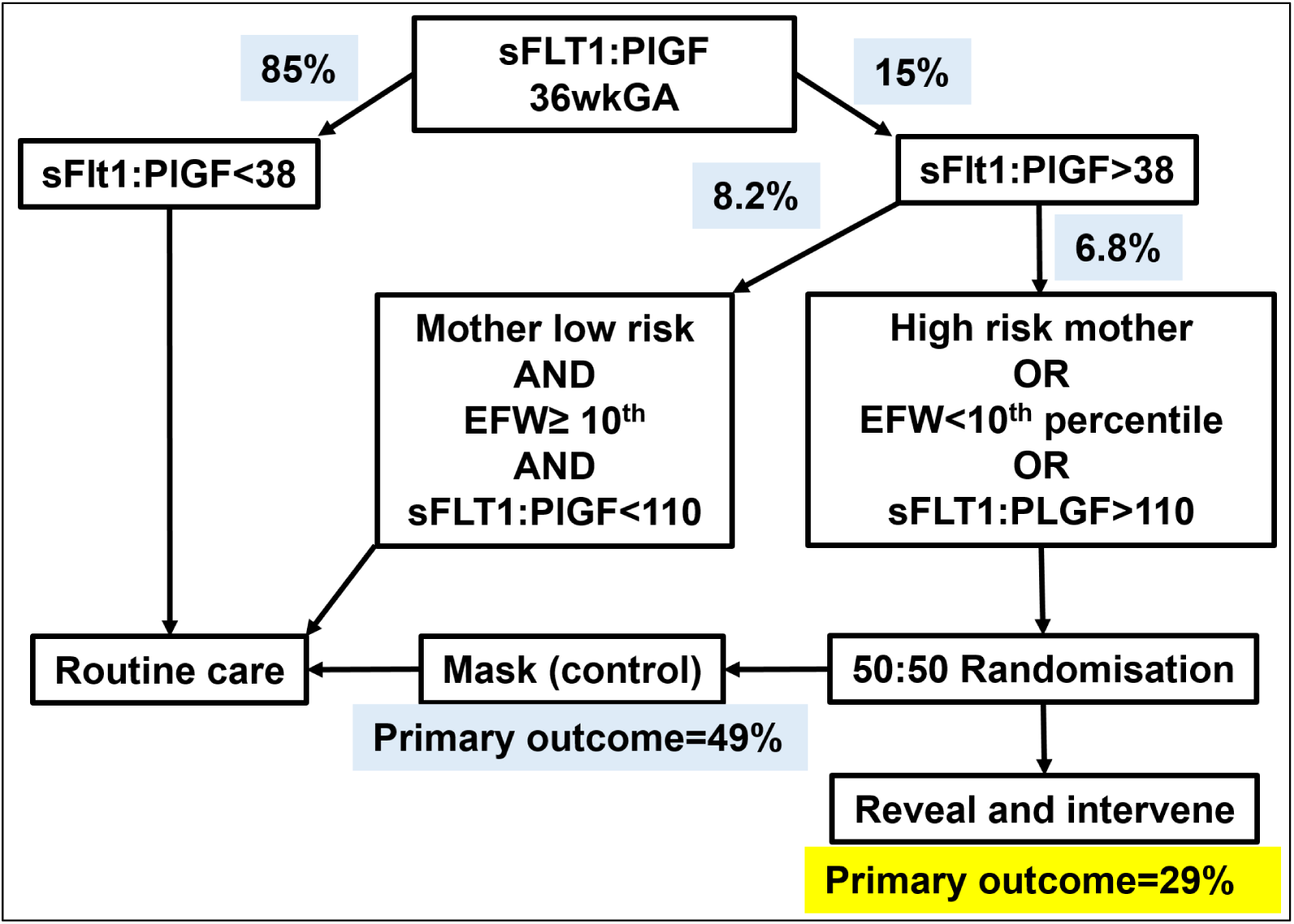
Flow diagram of risk assessment and randomisation. Percentages in blue boxes indicate the observed proportion in the original POP study. The yellow box indicates an estimated risk of the primary outcome among the women who screen high-risk and are offered induction of labour.

**Table 1.**
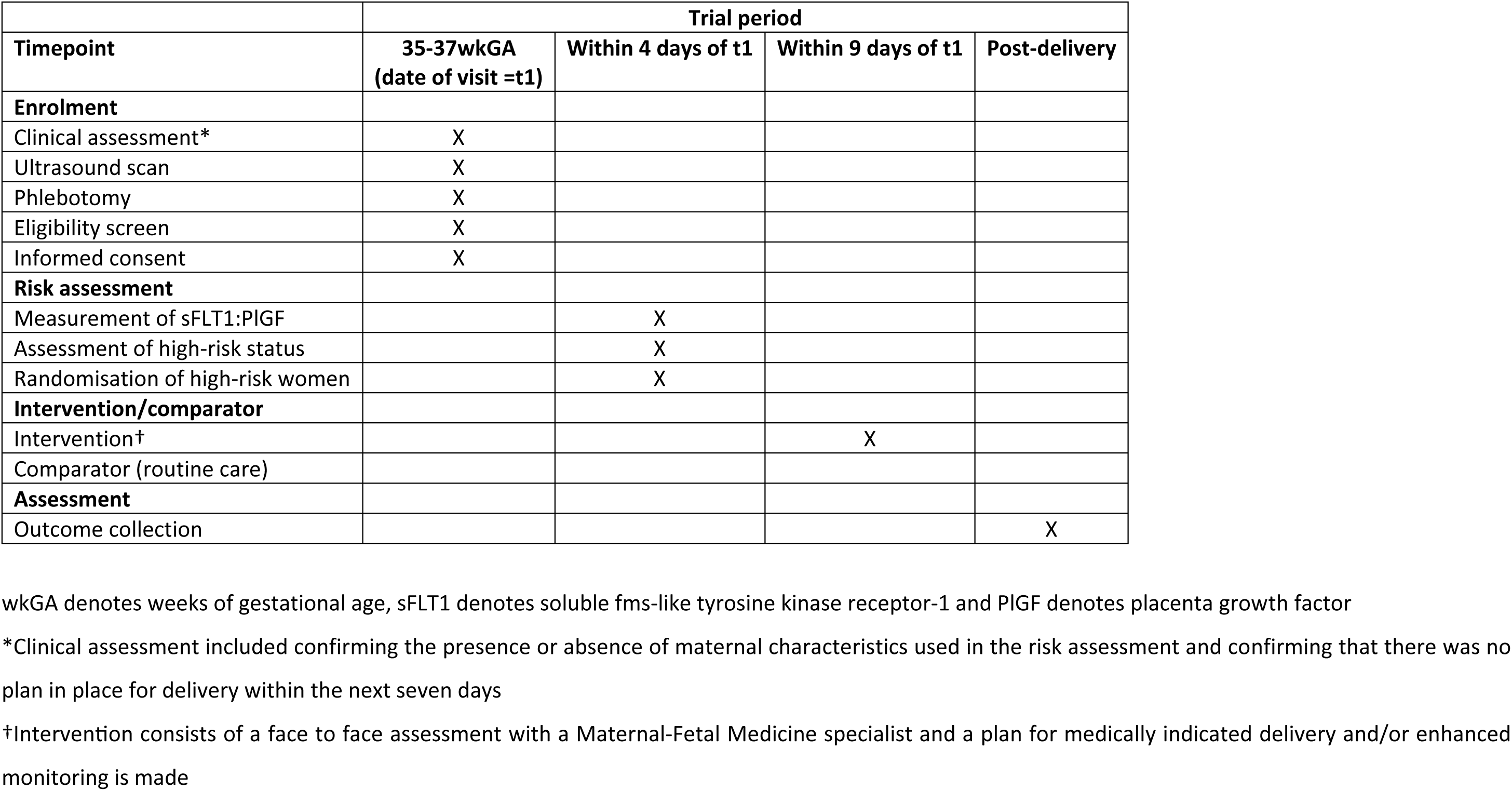
Schedule of enrolment, interventions and assessments for the RCT.

#### Visit 1 at dating scan

Fetal measurements made at the time of the dating scan will be recorded as well as the estimated date of delivery, using the Rosie Hospital guideline for gestational dating. Additionally, the participant will be weighed and the value (kg) will be recorded, she will be asked for her pre-pregnancy weight, and her height will be measured and the value recorded (cm). She will then be directed to the Clinical Research Facility, where bloods will be taken and appointments made for the next visit. Research bloods (<50ml in total) will be obtained, for example for serum, plasma, and cells. Booking bloods/Down’s syndrome screening bloods can also be performed if required, providing the woman has consented to screening as per routine clinical care.

#### Visit 2 at ∼20wkGA anomaly scan

Women will attend the Clinical Research Facility at 20wkGA for a research scan. The routine anomaly scan will be conducted as per local policies and procedures with referral for any concerns as required. It should be performed no earlier than 18 weeks + 0 days and no later than 20 weeks and 6 days. Any adverse findings at the anomaly scan should follow the Rosie referral pathway, e.g. to the Fetal Medicine Unit. The anomaly scan involves fetal biometry which will be revealed and any abnormalities actioned as per hospital protocols. The only modifications to the scan will be that the sonographer will also perform umbilical artery Doppler flow velocimetry (hereinafter referred to as “Doppler”) and uterine artery Doppler (both left and right), additional measurements will be made of the fetal head (biparietal diameter and the transthalamic head circumference), and the maximum vertical pool of amniotic fluid will be recorded. The following additional data will be recorded on the study database:

1. Right uterine artery Doppler pulsatility index
2. Presence of notch in right uterine artery Doppler
3. Left uterine artery Doppler pulsatility index
4. Presence of notch in left uterine artery Doppler
5. The mean uterine artery Doppler pulsatility index
6. Umbilical artery Doppler pulsatility index
7. Pattern of umbilical artery end diastolic flow (reversed, absent, forward)
8. Biparietal diameter
9. Head circumference (transthalamic section)
10. Maximum vertical pool of amniotic fluid

The results of umbilical artery Doppler will not be revealed. Uterine artery Doppler results will only be revealed if the patient has characteristics which require the performance of uterine artery Doppler as described in the NHS England Saving Babies’ Lives Care Bundle(14). At the same visit, basic clinical and demographic data will be obtained and participants will also have their weight measured. The results of both will be recorded in the study database.

#### 7.3 Visit 3 at ∼28wkGA

Women will attend the Clinical Research Facility at around 28wkGA for a research scan. These scans will not replace any scans which are clinically indicated. The following measurement will be made:

1. Right uterine artery Doppler pulsatility index
2. Presence of notch in right uterine artery Doppler
3. Left uterine artery Doppler pulsatility index
4. The mean uterine artery Doppler pulsatility index
5. Presence of notch in left uterine artery Doppler
6. Umbilical artery Doppler pulsatility index
7. Middle cerebral artery Doppler pulsatility index
8. Pattern of umbilical artery end diastolic flow (reversed, absent, forward)
9. Biparietal diameter
10. Head circumference
11. Abdominal circumference
12. Femur length
13. Placental localisation (anterior, posterior, left lateral, right lateral, fundal)
14. Placenta low lying (yes/no)
15. Fetal presentation (cephalic, breech, transverse, other)
16. Amniotic Fluid Index (AFI, i.e. the sum of the deepest pools of amniotic fluid in each of the four quadrants of the uterus)

These measurements will be recorded on the research database. Digital images will be saved. The information from these scans will not be recorded on the Astraia database (the hospital’s clinical obstetric ultrasound database), the patient’s EMR (EPIC), or in the hand-held notes and participants will not be informed of the full findings of the scan unless any of the following are observed:

- Intra-uterine fetal death
- AFI <5cm
- Any major fetal anomaly abnormality not previously noted
- Previously undiagnosed low lying placenta
- Persistently absent or reversed end diastolic flow in the umbilical artery

In the event that any of the above are observed, the woman will be informed, a full report will be printed and also placed on her EMR, and she will be referred to Clinic 22 (Maternal Medicine), Clinic 23 (Triage) or the Fetal Medicine Unit, as appropriate, for further assessment. If none of the above are observed, a limited Astraia report will be printed and also placed on her EMR which states that the findings above were not observed. This will clarify that fetal biometry, AFI and Doppler were not reported and it should not be assumed that they were normal. Hence, participants should continue to be referred for clinically indicated scans as per the Rosie Hospital’s guidelines. At the same visit, participants will also have their weight measured and the result recorded in the study database, and blood will be obtained (<50ml total).

#### 7.4 Visit 4 at ∼36wkGA

Women will attend the Clinical Research Facility at around 36wkGA for a research scan. These scans will not replace any scans which are clinically indicated unless the only indication for a scan is to determine presentation. The same measurements will be made as described above at the 28wkGA visit, and the same processes will be followed in relation to recording the findings. Participants will not be informed of the full findings of the scan unless any of the following are observed:

- Intra-uterine fetal death
- AFI <5cm
- Any fetal anomaly not previously noted
- Previously undiagnosed low lying placenta
- Persistently absent or reversed end diastolic flow in the umbilical artery
- Non-cephalic presentation

In the event that any of the above are observed, the woman will be informed, a full Astraia report will be printed and also placed on her EMR and she will be referred to Clinic 22 (Maternal Medicine), Clinic 23 (Triage) or the Fetal Medicine Unit, as appropriate, for further assessment.

If none of the above are observed, a limited Astraia report will be printed and also placed on her EMR which states that the findings above were not observed. This will clarify that fetal biometry, AFI and Dopplers were not reported and it must not be assumed that they were normal. If the woman, her midwife or doctor has any concerns in relation to fetal growth and well-being or any other aspect of the pregnancy that would normally result in referral for an ultrasound scan that a referral for a clinical scan will be made to the main ultrasound department. At the same visit, participants will also have their weight measured and the result recorded in the study database, and blood will be obtained (<50ml total).

#### Delivery

At the time of delivery, the attending midwife will try to obtain cord blood (arterial and venous) for storage and for blood gas analysis (this will not be possible in all cases). This is normally performed in cases of suspected fetal distress but will be performed routinely for all women in the study even in the absence of suspected fetal distress. The placenta, membranes and umbilical cord will be placed in a dedicated refrigerator in the clinical area. Technicians will attend regularly to obtain samples from the placenta and umbilical cord as previously described(7), and the placenta will subsequently be discarded as per standard procedures.

### Sample size

The sample size calculation was based on the observed proportions of the primary composite outcome and the timing of delivery in the POP study in the women who screened high-risk. There were 256 women who screened high-risk and 126 (49%) of them experienced the primary outcome. The predicted effect of induction of labour to prevent cases of the primary composite outcome differed according to whether the recommendation was induction of labour at 37 weeks or at 39 weeks. For the women where the criteria for 37 week induction were present, it was assumed that 25% of cases of preeclampsia before 38 weeks would be prevented, that 50% of cases of preeclampsia at 38 weeks would be prevented and that 75% of cases of preeclampsia at 39 weeks and beyond would be prevented, and it was assumed that the proportional reduction would be the same for cases where there was also neonatal morbidity or mortality. However, for preeclampsia plus fetal growth restriction, it was assumed that, overall, 40% of cases would be prevented, and the same assumption was made for fetal growth restriction without preeclampsia. This is a conservative estimate, as the DIGITAT RCT demonstrated a 60% reduction in the proportion of babies <3^rd^ percentile (from 30.6% to 12.5%) with a policy of induction of labour for ultrasonically suspected small for gestational age at 37 weeks(15). It was assumed that induction of labour would have no effect on neonatal morbidity or mortality in the absence of preeclampsia or fetal growth restriction. For the women where the criteria for 39wkGA induction of labour were present, it was assumed that 25% of cases of preeclampsia at 39 weeks and 75% of cases at or after 40 weeks would be prevented. It was conservatively assumed that induction of labour at 39 weeks would have no effect on birth weight <3^rd^ percentile or neonatal morbidity or mortality. Finally, as the interval between recruitment to the RCT and delivery will be about four weeks, we anticipate zero loss to follow up. Applying these assumptions to the POP study data indicated that 51 out of the 126 events would have been prevented, yielding a relative risk (RR) of ∼0.6. A sample size calculation indicates that we would need 246 high-risk women, allocated 1:1 to intervention or control, to have 90% power to detect a reduction in the primary outcome from 49% to 29% with a two-sided P value of <0.05. In the POP study, 6.8% of women screened positive hence we would need to recruit ∼3,600 women at 36 weeks to obtain this number of high-risk women. If we assume that 80% of women recruited to the cohort study would be eligible and would consent for the RCT, we calculate that we need to recruit 4,500 women. Given the uncertainty about the proportion who would be eligible for and would consent to the RCT, we recalculated on the basis of 60% of 4,500 women being eligible and consenting. This would result in an estimated 184 high-risk participants and would yield ∼80% power to identify the same proportional reduction.

The assumptions of the sample size calculation were evaluated prior to the 2024 meeting of the Trial Steering Committee (TSC). A report on recruitment was provided to the TSC. At the end of May 2024, 3667 women had been recruited to the cohort and this was as predicted, with an estimated ∼4,500 participants by the then planned time of cessation of cohort recruitment (end May 2025). However, the proportion of women who were eligible for the RCT and consented was 75% rather than the predicted 80%. Moreover, of the women who were recruited to the RCT, the proportion who screened high-risk was 6.2% rather than the predicted 6.8%. The result was that the predicted number of high-risk women in the RCT would be 195-200. It was estimated that if recruitment was continued for a further year, that ∼RCT500 women would be recruited to the cohort, ∼3,900 would be recruited to the RCT and 240-250 high-risk women would be randomised to intervention or control. The TSC unanimously recommended prolonging recruitment to the RCT and ethical approval was obtained for the new sample size.

The above ignores the lower P value required for the two primary outcomes. The method for adjusting the P value for the two primary outcomes is described in the associated statistical analysis plan. The actual P value threshold will depend on the degree of correlation between the two primary outcomes and this will only be known once all of the data have been collected. However, the adjusted P value threshold will be somewhere between 0.025 (calculated using a simple Bonferroni correction and assuming no correlation) and 0.05 (the threshold if there was only a single outcome). Using the conservative Bonferroni corrected threshold of 0.025, a study of 250 high risk women would have 85% power to detect a reduction in the primary outcome from 0.49 to 0.29. Hence the study should have a least 85% power to detect the expected reduction in the first primary outcome.

### Recruitment

The Patient Information Leaflet will be sent out with the appointment for the patient’s dating ultrasound. Research staff will liaise closely with the administrative team in the ultrasound department to ensure that this is taking place and will monitor the use of forms in the department. The initial approach about the study will be made by the sonographer performing the scan and the research team closely interact with the sonographers to remind them about the study and to address any barriers. Posters in the ultrasound department will also highlight the study to women attending and give the contact details for the recruitment team. Timing recruitment of fathers to the 20 weeks scan increases the likelihood of being able to approach them. As it will typically involve sharing information about the baby’s sex, this scan is probably the most highly attended of a patient’s ultrasounds. The initial approach for the second consent for the RCT will be made by the research sonographers and there will be very close interaction between this team and the research midwives obtaining consent to identify and overcome barriers to recruitment.

### Assignment of interventions: allocation and sequence generation

Allocation will be performed by block randomisation with an allocation ratio for intervention (reveal and offer early delivery) to control (do not reveal and participant has routine maternity care) of 1:1. The investigators are currently blinded to block size but this will be reported in the publication of the trial results. Randomisation will be stratified across the following groups:

1. sFLT1:PlGF ratio >110
2. sFLT1:PlGF ratio >38 + 36wkGA ultrasonic estimated fetal weight (EFW) <10th percentile
3. sFLT1:PlGF ratio >38 + 20wkGA uterine artery Doppler pulsatility index >90th percentile, or
4. sFLT1:PlGF ratio >38 + increased risk of preeclampsia using an adapted version of the NICE Guideline.

Where an individual falls into more than one of the above groups, allocation to a stratum will be based on the hierarchy above. e.g. a woman with a sFLT1:PlGF ratio of 120 and an EFW <10^th^ percentile would fall in stratum 1. Randomisation lists for each of the strata will be generated using Stata version 16.1 (StataCorp, College Station, TX, USA).

### Concealment mechanism

The randomisation process will be built into the study database. Where women fulfil the eligibility criteria there will be a separate randomisation screen and the allocation will be revealed by clicking a button on the screen. The database will be created by the MRC Cambridge Epidemiology & Trials Unit (CETU) and the investigators will have no control over the randomisation process and have no way of knowing the allocation prior to clicking the randomisation button. In the event that the computer system is unavailable, other methods of random allocation will be utilised, as indicated by the CETU. As this will be performed without stratification, the randomisation algorithm will be amended by an unblinded member of the statistical team when the system is available to maintain stratification across all women randomised in the study.

### Implementation

The allocation sequence will be generated by the CETU. Participants will be given a Patient Information Leaflet at the time of their 28wkGA research appointment. When participants attend the 36wkGA appointment, those who meet the inclusion criteria for the RCT element of the study will be asked by the sonographer performing the research scan if they are happy to talk with a research midwife. The consent process is described above. The Study Coordinator or a deputy will check the risk status of all women who appear to have screened high-risk based on the sFLT1:PlGF ratio and other factors. If these checks confirm the high-risk status, randomisation will be performed as described above. Women who are randomised to intervention will be contacted and a clinical visit will be booked to discuss their risk status and plan their subsequent care. High-risk women who are allocated to control will not be contacted and will receive care as per the clinical guidelines of the Rosie Hospital. The identity of screen positive control women will only be known by the individual performing randomisation and this individual will never be involved in the delivery of clinical care to that participant.

### Assignment of interventions: Blinding

Among the women who screen high-risk, those who are randomised to intervention will be known to the investigators and the clinical team as the patient’s care will be subsequently determined by the fact that they screened high-risk. The team member performing the randomisation will be the only individual who will know the identity of high-risk women randomised to routine care and they will not share that information with either the other members of the study team or clinicians in the hospital. The unblinded Trial Statistician, the CETU IT specialists and the Data Safety Monitoring Committee will be the only individuals with access to the identity of the high-risk women randomised to routine care. Similarly, when the investigators obtain extracts of the data (e.g. for the purposes of data cleaning), there will be nothing to identify high-risk women randomised to the control group.

### Procedure for unblinding if needed

The identify of women according to their randomisation status will be unblinded for all women randomised to intervention. The only individuals who will review data where the randomisation status will be fully unblinded are the unblinded Trial Statistician and the members of the Data Safety Monitoring Committee. There is no procedure for emergency unblinding as all of the data collected are for the purposes of research only. Low risk women and high-risk women randomised to control care will receive the normal standard of care for women in the Rosie Hospital and there is no requirement to access their research data and randomisation status in order to deliver that care.

### Data collection and management: plans for assessment and collection of outcomes

Collection of research data in the antenatal period will be performed at each of the four study visits, described above. All data will be entered directly onto a custom built database which will be prepared and managed by the MRC CETU. Data will be reviewed regularly, typically every month, and plotted to allow identification of outliers. Where an outlier is identified, the validity of the measurement will be checked from the original source. In the case of ultrasonic measurements, this will be achieved by comparison with the stored images obtained at the time of the scan. Collection of maternal demographic and medical data will be performed at the second visit. Outcome data will be collected by review of the EMRs, namely, EPIC for the mother and Badgernet and EPIC for the infant. Data will be entered onto paper forms and these will be sent to a company for transcription into an electronic format which employs double entry to reduce transcription errors. Again, on receipt of the data, it will be assessed for outliers and discrepant values and errors will be corrected by checking against the source document.

Determination of of the sFLT1:PlGF ratio will be performed using a Roche Cobas e411 automated analyser. The machine will be operated, serviced and calibrated according to the manufacturer’s instructions. All staff using the system will be experienced laboratory technicians who have received training on how to operate it. All staff using the machine and reporting results will have been certified for Good Laboratory Practice training and have refresher courses at recommended intervals. The threshold value of sFLT1:PlGF to indicate elevation is >38. All samples which yield an elevated value will be re-measured and the result will be only accepted if there is a second value which is within 10% of the first. The reported intra-assay coefficient of variation for human serum samples is <2% for sFLT1 and PlGF, and the inter-assay coefficients of variation are 2.3% to 4.3% for the sFLT1 assay and 2.7% to 4.1% for the PlGF assay(10).

### Plans to promote participant retention and complete follow-up

Outcome data will be collected from all participants, irrespective of any deviation from the intervention protocol. For women who screen low risk or who screen high-risk and are randomised to routine care, there will be no further study visits after the 36wkGA visit. All women will deliver their baby within eight weeks of this visit, hence we do not anticipate substantial loss to follow up. If women choose to deliver in a different hospital, we will liaise with the hospital in question to retrieve their outcome data. Women who screen high-risk and are randomised to intervention will typically be offered a face to face consultation with a consultant in Maternal-Fetal Medicine within a week of their 36wkGA visit. On-going care will be informed by a Clinical Guideline for high-risk participants, which has been incorporated into the hospital’s Clinical Guidelines database. On-going care will be determined in a process of shared decision-making between the consultant and the high-risk participant, who will be able to choose to accept or decline elements of care, such as early delivery or enhanced maternal and fetal monitoring.

### Data management

Outcome data will be collected by multiple methods. Research staff will access the participant’s records on EPIC to obtain outcome data. Information will be entered onto a standardised paper form and paper records will be subsequently transferred to a research database. Transcription of data into an electronic format involves will use double entry with checking of anomalous values. The data will be intermittently reviewed to assess completeness and to identify outliers, which will then be checked. Outcome data will also be collected by downloading the electronic data from participants from the different software packages used by CUHFT to store clinical information. This may include download of information from the main EMR, EPIC (e.g. laboratory records, vital signs, point of care testing, and notes for free text searching), ultrasound data (Astraia), and neonatal outcome data (Badgernet). Outcome data will be obtained from external Trusts in the event that a woman transfers her care after 36wkGA. Long term outcomes will be ascertained through linkage of the mother’s or baby’s records (e.g. for outcome of subsequent pregnancy, future inpatient or outpatient care [mother and baby], and educational achievement [baby only]). Examples of external data sources include NHS Digital, Public Health England and the Department for Education.

### Confidentiality

All data will be stored securely. Data obtained from the research visits will either be entered directly into an electronic Case Report Form (eCRF) and stored in a research database or data from paper CRFs can be entered into the database directly, for example, when access is suspended for technical reasons and data cannot be entered at the time of the visit. Hard copies of consent forms and paper copies containing data will be stored securely in a locked filing cabinet and will be securely destroyed when no longer required. Consent forms will be also scanned into PDF format and stored on the patient’s EMR. The research database for the eCRF will be accessed through CUHFT computers. The eCRF data from participants will be transferred to Cambridge University IT systems directly into a secure space, currently, the University of Cambridge Secure Research Computing Platform. This provides a password and firewall protected Safe Haven for members of the University to store sensitive data, including Personally Identifiable Data.

Electronic files of research data will be combined with electronic files of clinical data using the mother’s identifiers to link the databases. All data which includes maternal identifiers will be stored securely, using the methods described above. Extracts of the data may be passed on to third parties, such as research collaborators. Whenever data is passed on, all identifiers will be replaced with a unique coded identifier to preserve anonymity. The key linking this number to the participant identifiers will be held securely, as described above. It will be impossible for external collaborators to identify individuals on the basis of the data. Moreover, all transfers of data will require completion of a Data Transfer Agreement. This requires the recipient of the data to adhere to a series of conditions in relation to the anonymised extract and an appropriately authorised signatory from the receiving institution.

None of the research information will be fed back to participants, other than as described above (the absence of reveal features from the scan, ultrasound findings which fulfil the criteria for being revealed and, for women who participate in the RCT, screen high-risk and are randomised to intervention, the sFLT1:PlGF assay result and relevant scan results).

### Plans for collection, laboratory evaluation and storage of biological specimens for genetic or molecular analysis in this trial/future use

Blood sampling will generally be performed in the Clinical Research Facility. Samples will be processed according to Standard Operating Procedures. Serum and plasma will be placed immediately in a freezer in the Clinical Research Facility and will be transferred for longer term storage in the Department of Obstetrics & Gynaecology laboratories at -80°C. Sample tubes will be labelled with the hospital Case Record Number and the participant’s initials and date of birth, to minimise the risk of identification error. The freezers storing the samples will be locked and will be placed in areas where access is limited (by locks and/or badge entry systems) to authorised personnel only. Samples may be passed on to third parties. In this event, tubes will have all identifiers removed and the sample will be given a unique coded identifier. The code linking the sample to the identifiers of the given participant will be stored securely, as described for other patient identifiable data. The samples will be tracked using sample inventory and tracking software.

The sFLT1:PlGF ratio will be measured on maternal serum samples using the Roche Diagnostics Ltd Cobas e411 assay platform. This is a CE marked clinical grade analysis platform. The machine will be used and maintained according to the manufacturer’s recommendations. All staff operating the system have been appropriately trained by Roche Diagnostics Ltd and have also undergone Good Laboratory Practice training with appropriately scheduled refresher training.

The cohort study involves extensive collection of biological samples. Only maternal serum from the 36wkGA visit will be analysed shortly after collection as part of the RCT. The other samples will be stored for future analyses. Participants will be informed that we cannot specify all analyses which might be performed using their samples and that they are giving their samples as a “gift” and that the investigators can use the samples in any way with the aim of better understanding or predicting complicated and healthy pregnancy outcomes.

Some analyses of the biological samples may take place while the trial is still in progress. In analyses of samples prior to final unblinding, no information will be provided about which samples were obtained from high-risk women randomised to routine care.

### Statistical methods for primary and secondary outcomes

The statistical methods are described in the Statistical Analysis Plan (SAP), which is provided as Additional File 1 of the current paper. In brief, maternal characteristics will be summarised by proportions (%) for categorical and ordinal outcomes and by median (inter-quartile range) for continuous outcomes. The timing of birth (spontaneous and medically indicated) and delivery with preeclampsia will be plotted as cumulative incidence. To assess the screening performance of the test, women who screen positive and are randomized to intervention will be excluded and the screen positive control group will be weighted by the inverse of the sampling fraction of all screen positive women (weighting will equal two if the two randomised groups are identical in size). Screening performance will then be summarized using standard screening descriptive statistics.

To assess the effectiveness of the intervention, the RR (95% confidence intervals, CI) of the given outcome will be calculated in the screen positive intervention group, referent to the screen positive control group, using a Poisson model with robust standard errors(16), adjusting for the stratification criteria as categorical variables and other covariates associated with the primary outcome in the POPS cohort (see SAP for details). All analyses will be pre-specified in the SAP.

### Interim analyses

Interim analyses will assess efficacy and futility using predefined criteria, and will be performed annually by the unblinded Trial Statistician who will share the results with the Data and Safety Monitoring Committee (DSMC). The DSMC will also perform intermittent reviews of the number of serious adverse events. These analyses will be confined to the women who had screened positive and had been randomised to routine care or intervention, as the other women who consented for the trial and had screened low risk could not have had their care modified by participating in the trial. The exact terms for stopping will be defined in the DSMC charter, following agreement with the chair. The DSMC will also be able to propose early cessation if analysis of severe adverse events or unexpected severe adverse events led them to conclude that the continuation of the trial was unsafe. In this event recruitment to the non-interventional element of the study would be continued until 5,500 women had been recruited. The DSMC will make recommendations about cessation but the ultimate decision to continue or to stop the trials will lie with the Trial Steering Committee (TSC).

### Methods for additional analyses (e.g. subgroup analyses)

These methods are described in the SAP. In brief, the screening performance of the test will be compared across the four categories of screen positive by plotting the positive predictive value and 95% CI for each group, and a Chi squared test will be performed to determine whether any apparent variation in the strength of prediction was greater than could be expected by the play of chance. The effectiveness of the intervention will be compared across the four categories of screen positive by plotting the adjusted RR and 95% CI associated with intervention, and a test for interaction will be performed using the Poisson model to determine whether any apparent variation in the effectiveness of the intervention was greater than could be expected by the play of chance.

### Statistical methods to handle protocol non-adherence and missing data

The analysis will be by intention to treat, irrespective of whether women in the intervention arm accepted the recommended timing of early delivery. Missing outcome data will be handled using complete case analysis and missing baseline covariate data will be handled by mean imputation in the multivariate analysis. If there is a >5% imbalance in missing outcome data between randomised groups, we will also evaluate whether results are robust to multiple imputation under missing not at random assumptions.

### Plans to give access to the full protocol, participant level-data and statistical code

The study protocol is described in the current paper, which is open access. Participant level data will not be publicly available as this is outside the ethical permissions for the study. However, anonymised data may be shared on request to the PI with appropriately qualified researchers through a Data Transfer Agreement, which would require an appropriately authorised signatory from the receiving institution. The statistical code for the primary analysis will be shared on request.

### Oversight and monitoring

#### Composition of the coordinating centre and trial steering committee

Day to day operational running of the trial will be led by the Study Coordinator and overseen by a Trial Management Group (TMG), chaired by the Chief Investigator, which will meet approximately every four weeks. The TMG will include the Study Coordinator, the Lead Sonographer, the head laboratory technician, the scientific lead for the laboratory, the Trial Statistician, and a representative from the CETU. A smaller sub-group, consisting of the Chief Investigator, the Study Coordinator and the Lead Sonographer, will be convened between these meetings. Clinical issues arising from the trial will be managed by the research midwives and research sonographers, with oversight by sub-specialist consultants in Maternal-Fetal Medicine.

The TSC will meet at least annually while the study is recruiting and will consist of an independent chair (an experienced perinatal trialist), an independent obstetrician, an independent neonatologist, an independent statistician, the Chief Investigator, and a lay representative. The role of the TSC is to provide overall supervision and to ensure that the trial is conducted to the appropriate standards, and these will be defined in the TSC Charter.

Monitoring oversight of the trial will be managed by the CETU. All elements of the oversight will conform to the CETU standard operating procedures, including working practices, case report forms, data management, and statistical analysis. CETU staff (database programmer, data manager and statistician) and the Trial Statistician will manage data generated by the trial.

#### Composition of the data monitoring committee, its role and reporting structure

The Data Safety Monitoring Committee (DSMC) will meet at least annually while patients are being randomised to intervention or control and will consist of a chair (an experienced perinatal trialist), an obstetrician (this role could also be the chair), a neonatologist, and a statistician, all of them independent from the study and the Rosie Hospital. All members of the DSMC will have prior clinical trials experience and no direct and relevant conflicts of interest. The roles and responsibilities of the DSMC will be defined in the DSMC Charter and will include (a) to review the un-blinded data comparing high-risk women randomised to intervention versus routine care, (b) to use the data to perform an interim analysis for safety, futility and efficacy, (c) to review unexpected serious adverse events among high-risk women as they occur, and (d) to perform intermittent comparisons of serious adverse events among high-risk women, comparing the control and intervention groups.

#### Adverse event reporting and harms

Adverse events will be defined as an untoward medical occurrence experienced by a trial participant. The majority of nulliparous women will experience an event that fulfils the definition of an adverse event around the time of delivery, such as perineal trauma, operative vaginal delivery or caesarean section. Serious adverse events will be defined as ≥1 of the following: (a) lethal, (b) life threatening, (c) requiring hospitalisation or prolonging hospitalisation, (d) resulting in significant or persistent disability or incapacity, (v) congenital anomaly or birth defect. Again, serious adverse events will be common among nulliparous women. For example, in the original POP study, 52% of the population fulfilled the above definition by virtue of experiencing an operative delivery (vaginal or caesarean), given that operative delivery can result in a prolonged length of stay in hospital. Serious adverse events for both mother and baby will be recorded for the trial and the relevant serious adverse events are captured by the list of primary and secondary outcomes.

The vast majority of serious adverse events in the study will be expected and will not be reported. However, we will report unexpected serious adverse events occurring to high-risk women who were randomised to control or intervention groups, which will be defined as per a previous RCT of pregnant women with preeclampsia comparing planned delivery versus expectant management in late pregnancy, the Phoenix Trial (ISRCTN01879376). The definition employed in that trial was one or more of the following: (a) maternal death, (b) maternal stroke, (c) stillbirth, (d) neonatal death. Each case will be reviewed by the Chief Investigator and by the local clinical risk management process. If either review indicates a likely causal link between the event and participation in the trial, the details will be referred to the DSMC for review within 14 days of the information coming to light. Unexpected serious adverse events will be reported to the Sponsor within 24h of research staff becoming aware of the event, and it will also be reported to the Research Ethics Committee (REC) which gave a favourable opinion to allow assessment of whether it was related in any way to the research procedures. Unexpected serious adverse events will also be reviewed by the DSMC during the conduct of the study.

#### Frequency and plans for auditing trial conduct

Monitoring oversight of the trial will be managed by the MRC CETU which is independent of the Chief Investigator and sponsor. All elements of the oversight will conform to the CETU standard operating procedures, including working practices, case report forms, data management, and statistical analysis. The format of the report will include a summary table of action points which will be reviewed by the TMG, with the Study Coordinator leading on providing responses to the report.

#### Plans for communicating important protocol amendments to relevant parties (e.g. trial participants, ethical committees)

Amendments to the study protocol will be communicated to the ethics committee via the submission of a major amendment, and this is managed by the UK’s Integrated Research Application System (IRAS). This process also requires review and approval by the sponsor, hence the information will be also reviewed by the CUHFT R&D Department. The TSC and DMC will be informed of any protocol changes at the time of their annual meeting. Very substantial changes in the protocol would be communicated to the members of these committees by email if there was not an imminent committee meeting planned. Any changes to the protocol would be communicated to participants through the Patient Information Leaflet and the Consent Forms. Changes in these documents would also be approved by the ethics committee as part of a major amendment on IRAS.

#### Dissemination plans

The primary mode of dissemination of the results of the trial will be though publication in a peer reviewed journal. We also anticipate presenting the results of the study at an international conference. The paper will be Open Access to allow it to be read by the general public. We will promote dissemination of the results through social media (X and Facebook) and by posting the results on the study website with a link to the full paper. We will liaise with the University of Cambridge press office regarding a press release to disseminate the results through news media, if appropriate. Authorship of study publications will be determined by ICMJE criteria and professional writers will not be employed.

## Discussion

A relatively unusual feature of the study is that it is large but is based in single centre. There were two elements to the rationale for the decision to confine the study to one centre. First, the Rosie Hospital is strongly supported by research infrastructure, in particular, the NIHR Cambridge Clinical Research Facility and the NIHR Cambridge Biomedical Research Centre. The existence of the infrastructure allows the required funding to be below the ceiling of the funder. Second, the Chief Investigator has been a consultant in Maternal-Fetal Medicine in the hospital for >20 years and we are confident that the process of screening and intervention can be implemented with high fidelity in the hospital. Studies may falsely conclude a negative result because the intervention was not fully and effectively implemented.

The population in Cambridge is relatively homogeneous and tends to be of above average socio-economic status. This has a number of implications for the data arising from the study. First, the results may not be generalisable to other populations with different socio-economic or ancestry profiles. However, the sFLT1:PlGF ratio has been shown to be predictive of preeclampsia in studies throughout the world(17). Moreover, the intervention of induction of labour has been widely evaluated in a large number of trials throughout the world in other contexts(18). Hence, despite the somewhat selected population, there is a high probability that a positive result would be replicable in other parts of the world. Second, because of the generally healthy state of the population, the prior risk of disease may be relatively low. This actually makes the study a more stringent evaluation of the novel screening approach. The positive predictive value associated with being screen positive is determined by the product of the prior odds of disease and the positive likelihood ratio associated with a positive screening result(19). Evaluating a screening test in a cohort with a low *a priori* risk means that the positive predictive value associated with screening positive will be lower than in the general population of pregnant women. It follows that, if the result of the trial is positive, it is very likely that the result will be replicable in other populations, as the proportion of true positive results could be even higher in the general population. However, conversely, a negative result would not rule out the utility of the screening approach in higher risk women. Finally, a major aim of the present study is to generate a biobank for future laboratory work to generate novel biomarkers and screening tests. Much of this work will employ “omic” methods in biomarker discovery, as we have described elsewhere(20). A major issue in the analysis of omics data from human clinical samples is separating signal from noise due to the very large number of hypothesis tests which these methods frequently generate. Measurements from a very heterogenous population will tend to have more noise than measurements taken from simple populations. The relatively homogeneous structure of the Cambridge population may be a positive factor when comparing “omic” data between cases and controls.

A further feature of the study which has implications for future implementation is the fact that women will not be randomised to being screened or not being screened but, rather, high-risk women will be randomised to having the result concealed or revealed. The advantages of this approach are summarised in detail elsewhere(6) but, in brief, it reduces the sample size required and, in the event of a negative result, allows us to determine whether screening failed because of failure of the screening test, failure of the intervention, or both (see Figure 1). However, the drawback of the design is that the trial does not directly simulate what a healthcare system could anticipate if the screening programme was introduced. For the reasons above, we consider the trial to be “proof of principle”, i.e. it is not certain that the result would lead directly to clinical implementation. A positive result may lead to a much larger, multicentre study where randomisation occurs at the point of screening or not screening. The current study should provide the data required for a sample size calculation. One potential study design which combines implementation and evaluation is a cluster, stepped wedge RCT and this may be an appropriate next step in the event of a positive result.

### Trial status

The current paper is based on version 3 of the study protocol which was finalised on 22^nd^ July 2024. However, the paper has been supplemented and reformatted for the purposes of creating a protocol paper, including compliance with the SPIRIT guidelines (21) and the journal’s Instructions to Authors.

Recruitment to the study commenced on 13^th^ January 2020 but was suspended on 17^th^ March 2020 due to the Covid-19 pandemic. Recruitment re-started on 3^rd^ August 2020. We currently anticipate recruitment to the cohort study will cease in June 2026, following the revision of the power calculation (see above), and recruitment to the RCT will cease approximately six months after recruitment to the cohort (Figure 1).

## Supporting information

Statistical analysis plan

SPIRIT checklist

## Data Availability

Participant level data will not be publicly available but may be shared with appropriately qualified researchers through a Data Transfer Agreement, which would require an appropriately authorised signatory from the receiving institution.

## Abbreviations

CETU: Clinical Epidemiology and Trials Unit
CI: confidence interval
DSMC: Data and Safety Monitoring Committee
EMR: Electronic Medical Record
IRAS: Integrated Research Application System
NICU: neonatal intensive care unit
PlGF: placenta growth factor
RCT: randomised controlled trial
RR: relative risk
SAP: statistical analysis plan
sFLT1: soluble fms-like tyrosine kinase receptor-1
SGA: small for gestational age
SCBU: special care baby unit
TSC: trial steering committee
wkGA: weeks of gestational age.

## Declarations

## Authors’ contributions

GCSS is the Chief Investigator, wrote the first draft of the protocol and SAP, and led the proposal and protocol development. DSCJ is the basic science lead. US is the unblinded statistician and both contributed to the study design. ASC is the Study Coordinator. ED is the lead ultrasonographer. EC is the head technician for sample management and analysis. IW is the blinded statistician. All authors contributed to the development of the protocol and approved the final manuscript. As US had seen unblinded data at the time of writing, she did not contribute to the writing of the SAP but she checked it for errors and factual content. All authors have read and approved the final manuscript.

## Funding

The primary source of funding for the present study is the Wellcome Trust (3,184,828 GBP). The study receives additional funding from Roche Diagnostics Ltd, who provide the resources required to perform the quantitation of sFlt1 and PlGF plus 100,000 GBP. Extracts from the contracts with Roche Diagnostics Ltd are provided as an additional file. The study also receives support from the NIHR Clinical Research Network, who provide salary support for research staff who recruit participants; the NIHR Cambridge Clinical Research Facility, which provide the rooms where the study visits take place and who provide supporting staff (nursing, administrative, phlebotomy and sample processing), and the Antenatal, Maternal and Child Health theme of the NIHR Cambridge Biomedical Research Centre, which funds core posts in the Department of Obstetrics and Gynaecology, University of Cambridge (UK), which support the study. IW was supported by the Medical Research Council Programme MC_UU_00004/07. None of the funders have contributed to or have authority over the design of the study, the collection, analysis, and interpretation of data or writing of manuscripts.

## Availability of data and materials

The Chief Investigator and Trial Statistician will have access to the final trial dataset. The primary point of access for the final trial dataset will be the Chief Investigator. Participant level data will not be publicly available but may be shared with appropriately qualified researchers through a Data Transfer Agreement, which would require an appropriately authorised signatory from the receiving institution.

## Ethics approval and consent to participate

The study has been approved by the East of England (Essex) Research Ethics Committee (19/EE/0331), the NHS Health Research Authority (IRAS271826) and the Cambridge University Hospital NHS Foundation Trust’s Research and Development Office (A095380). All participants will provide written, informed consent. The views expressed are those of the authors and not necessarily those of the NHS, the NIHR, or the Department of Health and Social Care.

## Consent for publication

The authors are willing to provide a model consent form on reasonable request.

## Competing interests

Within the scope of this work (ever):

GS and DSC-J have received funding from Roche Diagnostics Ltd to support the study. This consisted of 436,577 GBP of payment in kind (loan of machine, consumables, servicing and training) and funds (100,000 GBP) to support research expenses (equipment and consumables). GS department received payment from Roche Diagnostics Ltd for a talk given by GS on the use of the sFLT1:PlGF ratio to predict fetal growth restriction in 2018. Roche Diagnostics Ltd also provided consumables and equipment to the value of 596,142 GBP in 2014 to perform the analysis of sFLT1 and PlGF in the original POP study samples.

Outside the scope of this work (last three years):

GS and DSC-J have received research support from Illumina and Pfizer (fetal growth restriction and preeclampsia, preterm birth and infection). GS is/has been a paid consultant to GSK (preterm birth), Natera (biomarker discovery), Medicines360 (screening in pregnancy), and is/has been a member of Data Monitoring Committees trials of vaccination in pregnancy (GSK and Moderna) and a novel therapeutic for hyperemesis gravidarum (NGM Biopharmaceuticals). DSC-J is/has been a paid consultant to Medicines360 (screening in pregnancy) and GondolaBio (pregnancy related diagnostics and therapeutics). GS, DSC-J and US are named inventors in a series of patents for the prediction of fetal growth restriction, preeclampsia and gestational diabetes mellitus in pregnancy, filed by Cambridge Enterprise and licensed to Medicines360.

## Authors’ information (optional)

GCSS is a Professor and the Head of the Department, in the Department of Obstetrics and Gynaecology at the University of Cambridge (UK) and a consultant in Maternal-Fetal Medicine at the Rosie Hospital, Cambridge, and is the Chief Investigator of the study.

ASC is a research midwife working in the Department of Obstetrics and Gynaecology at the University of Cambridge (UK) and is the Study Coordinator.

ED is a research sonographer in the Department of Obstetrics and Gynaecology at the University of Cambridge (UK) and is the Lead Sonographer for the study.

US is a Research Professor in the Department of Obstetrics and Gynaecology at the University of Cambridge (UK) and is the undblinded Trial Statistician.

EC is a senior research laboratory technician and manages the team of laboratory technicians working on the trial.

IW is Professor of Statistical Methods for Medicine at University College London and is the blinded statistician for the study.

DSC-J is Professor of Reproductive Biology in the Department of Obstetrics and Gynaecology at the University of Cambridge (UK) and directs the laboratory elements of the study.

## Additional Files

1. Statistical Analysis Plan.
2. Completed SPIRIT+ checklist

## Notes

### Clinical Trial

ISRCTN12181427

### Author Declarations

The study has been approved by the East of England (Essex) Research Ethics Committee (19/EE/0331), the NHS Health Research Authority (IRAS271826) and the Cambridge University Hospital NHS Foundation Trust Research and Development Office (A095380).

