## Supplementary material for "A prospective cohort study of unselected nulliparous women with a nested randomized controlled trial of screening using the sFLT1:PlGF ratio, ultrasound and maternal characteristics and intervention using enhanced monitoring and early delivery: study protocol for the POPS2 cohort and randomized cont": Statistical analysis plan

### 1. Administrative information

**Title:** STATISTICAL ANALYSIS PLAN FOR THE POPS2 TRIAL

**Trial registration number:** ISRCTN 12181427

**SAP Version** 5

**Protocol Version** 4

**SAP Revisions**

| Version | Date | Description |
|---|---|---|
| 1 | 20/11/24 | First draft completed by GS |
| 2 | 22/10/25 | Second draft following revision by IW and discussion between IW and GS. US performed analysis of POPS cohort to identify variables to use in statistical adjustment of primary outcome |
| 3 | 11/12/25 | Third draft reformatted as per JAMA 2017 Guideline and incorporating further input from IW |
| 4 | 19/12/25 | Draft of final agreed version after further input from IW |
| 5 | 09/01/26 | Final agreed version after further minor edits by all authors |

**Roles and responsibilities:**

Gordon C S Smith, Chief Investigator. Professor of Obstetrics & Gynaecology, Department of Obstetrics & Gynaecology, University of Cambridge, UK.

Ulla Sovio, Unblinded Statistician. Research Professor in Biostatistics, Department of Obstetrics & Gynaecology, University of Cambridge, UK.

Ian R White, Blinded Statistician. Professor of Statistical Methods for Medicine, Medical Research Council Clinical Trials Unit at University College London, UK.

### 2. Introduction

2.1 Background, rationale and scope

This document describes the statistical analysis plan (SAP) for the Pregnancy Outcome Prediction 2 study (POPS2). The study is a follow up to the Pregnancy Outcome Prediction Study (POPS) which was conducted between 2008 and 2013 and is described elsewhere(1, 2). The SAP is provided as a supplement to the POPS2 protocol paper and the two documents should be read in conjunction. Readers are referred to the accompanying protocol paper for full details of the design and conduct of the trial. The SAP was written at around 80% of the recruitment to the trial and was finalised when recruitment was around 90%. Members of the Data Safety and Monitoring Committee had seen unblinded data as part of the trial monitoring process and did not contribute to the writing of the SAP due to their potential to be influenced by trends observed in their assessment of unblinded data prior to trial completion. The only exception to this is that the unblinded statistician, Dr Ulla Sovio, performed an analysis of the POPS cohort to identify which variables should be used in the multivariate analysis of the outcomes in the current study, and that analysis is included in the SAP.

The scope of the current SAP is for the primary analysis of POPS2 which will assess the screening performance of the test and the result of intervention in high-risk women. However, as described below, POPS2 also involves collection of extensive data and maternal blood, plus samples obtained at the time of delivery for clinical and translational research. Like other prospective cohort studies, both POPS and POPS2 are platform studies which, although prospective in conduct, facilitate retrospective analysis of data and biological samples. Analyses of these future studies are outside the scope of the current document.

2.2 Overview and objectives

POPS2 is a cohort study which recruits nulliparous women with a non-anomalous singleton fetus at the time of their dating ultrasound in the Rosie Hospital, Cambridge, United Kingdom, typically at around 12 weeks of gestational age (wkGA). Women have blood taken around the time of their dating ultrasound, following consent and recruitment, and at three subsequent research visits at around 20, 28 and 36 wkGA, which also include ultrasonography. At each visit, some elements of the ultrasound scan result are revealed and reported in the hospital's Electronic Medical Record (EMR, Epic) and some elements are not reported. Blood from the participant's partner is also obtained at one of the follow up visits. At the time of the 36wkGA visit women are asked for consent to participate in the RCT. All women who consent have blood taken to determine the sFLT1:PlGF ratio using a Roche Cobas e411 analytic platform. Women are assessed as high-risk on the basis of an extremely elevated sFLT1:PlGF, or an elevated sFLT1:PlGF ratio plus the presence of other risk factors. High-risk women are then randomised to having the result revealed or having the result concealed. When revealed, the participant is seen by a consultant in Maternal-Fetal Medicine and enhanced monitoring and/or early delivery is offered. Women who have the result concealed have the same clinical care that they would have experienced had they not participated in the study. The first primary outcome for both the screening study and the interventional study is a composite of perinatal morbidity and mortality and maternal experience of preeclampsia. The second primary outcome is a subset of the first and is defined as the most severe manifestations of perinatal morbidity and mortality and maternal experience of preeclampsia. The screening test was generated through analysis of the previous cohort study, POPS. POPS2 is designed to allow validation of the screening test (by comparing high-risk women randomised to routine care with low risk women) and evaluation of the effect of the intervention in high-risk women (by comparing high-risk women randomised to routine care and high-risk women randomised to intervention). Finally, both POPS and POPS2 involve collection of other biological samples (maternal blood, serum, plasma, paternal blood or saliva, placenta, umbilical cord, umbilical cord blood and fetal membranes) for the purposes of translational research outside the scope of the RCT.

**3. Study methods**

The full study methods are described in the accompanying protocol paper, including the trial design, randomisation, sample size, framework, statistical interim analysis and stopping guidance, timing of final analysis and the timing of outcome assessments.

**4. Statistical principles**

4.1 Confidence intervals and p values

The threshold for statistical significance is set at 0.0RCT two sided. In the analysis, the two primary outcomes will be treated as multiple primary endpoints, i.e. we will conclude a positive result if either of the endpoints is significantly less frequent in the intervention group compared with the control group. Treating the two outcomes in this way requires an adjustment to control the type 1 error rate at 0.05(3). We will correct the p value threshold for the use of two primary outcomes using the appropriate tail area of the bivariate normal distribution, which accounts for the correlation between the two primary outcomes, which we will estimate from the randomised data pooled across arms. No adjustment will be made for the number of comparisons in the secondary outcomes. If either of the two primary outcomes is significantly less common in the intervention group, any statistically significant differences in the secondary outcomes which were used to define the composite outcome will be interpreted as providing evidence that the intervention affected the likelihood of the given outcome.

4.2 Adherence and protocol deviations

Major non-compliance is defined as a case where the data are considered to be fraudulent.

Minor non-compliance will include:

- participants who are recruited to either the cohort or the RCT but who do not fulfil the relevant inclusion criteria
- participants who are recruited to either the cohort or the RCT who have one or more of the relevant exclusion criteria
- participants without fully documented consent for any element of the study
- participants who consented to the RCT but a reliable measurement of sFLT1:PlGF ratio was not obtained
- participants who are recruited to the RCT, screen high risk and are randomised to control where the blinded elements of the 36wkGA ultrasound scan or the sFLT1:PlGF ratio are revealed

Any cases with major non-compliance will be excluded from the analysis. Cases of minor non-compliance will be analysed by intention to treat if there is a valid assessment of their high-risk status according to the protocol and they are subsequently randomised. High-risk women who are randomised to the intervention group but decline any intervention are not regarded as cases of protocol non-compliance, as the study is evaluating the outcome of revealing the results of the screening test only. In the study, as in eventual clinical practice if the screening test was implemented, some women will decline intervention.

4.3 Analysis populations

There are four groups within the study:

(a) women who are recruited to the cohort study but are not recruited to the RCT

(b) women who are recruited to the cohort study and the RCT who screen low risk

(c) women who are recruited to the cohort study and the RCT who screen high-risk and are randomised to routine care

(d) women who are recruited to the cohort study and the RCT who screen high-risk and are randomised to intervention

The current analysis plan relates to groups (b), (c) and (d). Women in group (a) will have their data and samples analysed when studying the whole cohort, an area outside the scope of this SAP, other than in the presentation of their demographic and clinical data in Table 1.
The analysis of the screening assessment element of the present study will compare groups (b) and (c). The analysis of the interventional element of the present study will compare groups (c) and (d). The screening analysis will determine whether the screening test was effective in identifying women at increased risk of the primary outcomes and its components. The interventional analysis will determine whether intervening in high-risk women reduces the risk of the primary composite outcome and whether there is evidence that intervention affects any of the secondary outcomes. Hence, the current study is designed to allow both assessment of the diagnostic effectiveness of the screening test and determination of whether intervening in women who screen positive reduces the risk of the primary outcomes. In contrast, a study of screening which randomises women to being screened or not being screened only allows an overall assessment of the effect of screening and intervention. In the event of a negative result, it is not possible to know whether screening and intervention failed because the screening test did not correctly identify high-risk women, whether the intervention was ineffective at preventing the outcome, or both.

**5. Trial population**

5.1 Screening for eligibility
Women will be identified as potentially eligible for the study if they are attending for their dating ultrasound scan and they have not previously had a birth.

5.2 Eligibility
Women will be eligible to be asked to consent to the study if they are 16 years or older and have not previously had a birth >23 weeks of gestational age (wkGA), have an apparently healthy singleton fetus at the time of the dating scan, and fetal biometry indicates that the pregnancy is less than 18wkGA at the time of the scan.

5.3 Recruitment
The CONSORT flow diagram will indicate the number of women who were deemed potentially eligible, the number who declined to participate, and the number who provided written informed consent at the dating ultrasound scan, the number who withdrew or were lost to follow up, and the number participating in the RCT. These elements are documented below.

5.4 Withdrawal and loss to follow up

5.4.1 The cohort study

The average duration of participation in the study is about six months, and three additional study visits are planned after the initial recruitment visit. For each of the subsequent study visits (i.e. at ~20, ~28 and ~36wkGA) the CONSORT flow diagram will document the numbers of women between the given visit and the previous visit who experienced the following: withdrawal from the study, delivery or miscarriage prior to the study visit, loss to follow up, and did not attend the visit (either because they failed to attend the appointment or because there was no capacity to offer the appointment).

5.4.2 The randomised controlled trial (RCT)

The CONSORT flow diagram will identify the number of women who attended for the 36wkGA visit, the number who were eligible for the RCT, the number who consented for the RCT, the number who had a valid risk assessment, the number who screened high risk, and the numbers of high risk patients who were randomised to intervention or control.

5.5 Baseline patient characteristics

Table 1 indicates the baseline characteristics which will be compared between the different groups of patients in the study.

**Table 1.** Baseline characteristics for the POPS2 trial

| **Characteristic** | |
|---|---|
| Maternal age – year | |
| | Median |
| | IQR |
| | Maternal age ≥40 – no. (%) |
| Age stopped full-time education (FTE) – year | |
| | Median |
| | IQR |
| Race or ethnic group – no. (%) | |
| | White |
| | Black |
| | Asian |
| | Other |
| Married – no. (%) | |
| Previous pregnancy loss <12wks – no. (%) | |
| | One |
| | Two or more |
| Previous pregnancy loss 12-23wks – no. (%) | |

| | |
|---|---|
| | One |
| | Two or more |
| Smoking status – no. (%) | |
| | Never |
| | Quit pre-pregnancy |
| | Quit during pregnancy |
| | Current |
| Alcohol use – no. (%) | |
| | Never |
| | Quit pre-pregnancy |
| | Quit during pregnancy |
| | Current |
| Body mass index (BMI) in early pregnancy | |
| | Median |
| | IQR |
| | BMI ≥35 – no. (%) |
| Conception – no. (%) | |
| | IVF |
| | Other infertility treatment |
| | Spontaneous |
| Pre-pregnancy medical complications – no. (%) | |
| | Hypertension |
| | Diabetes mellitus (type 1 or 2) |
| | Chronic renal |
| | Immunological (SLE/APS) |
| 20wks uterine artery Doppler – mean pulsatility index | |
| | Median |
| | IQR |
| | >90$^{th}$ percentile |
| 36wks estimated fetal weight | |
| | Median |
| | IQR |
| | <10$^{th}$ percentile |
| sFLT1:PlGF | |
| | Median |
| | IQR |
| | >38 – no. (%) |

IQR denotes inter-quartile range, SLE denotes systemic lupus erythematosus, APS denotes anti-phospholipid syndrome

Continuous data will be summarised using the median and interquartile range, as these are informative irrespective of whether the data are normally or non-normally distributed. Categorical outcomes will be summarised as n (%). The demographic and obstetric characteristics of four groups of women will be tabulated: (a) women who were recruited to the cohort but did not consent to the trial, (b) women who were recruited to the trial and who screened low risk, (c) women who were recruited to the trial, screened high risk and were randomised to control, and (d) women who were recruited to the trial, screened high risk and were randomised to intervention. We will not employ statistical hypothesis tests to compare these characteristics between the four groups.

**6. Analysis**

6.1 Outcomes

As described in section 4.3, above, the study has two elements. The first is the analysis of the diagnostic effectiveness of the screening test. The second is the analysis of the clinical effectiveness of the intervention in women who screen high risk. The primary composite outcomes are the same for both elements. When analysing the diagnostic effectiveness of the screening test, data will only be presented on the primary composite outcomes and the individual elements of the composite outcomes, as the elements of the screening test were selected on the basis that they were associated with the components of the composite outcomes. For the analysis of the effects of intervention in patients who screen high risk, the analysis will also address the primary outcomes and the individual elements of the composite outcomes, but, additionally, a range of secondary outcomes will also be analysed. The rationale for studying the effects of the intervention on the secondary outcomes is that intervention in high-risk patients could cause other benefits or harms which are not necessarily predicted by the screening test.

6.1.1 Primary outcomes

Both primary outcomes are composites. The first is one or more of the following: (i) diagnosis of preeclampsia (using the ACOG 2013 definition)(4), (ii) perinatal death or perinatal morbidity, or (iii) delivery of an infant with a birth weight <3rd percentile for sex and gestational age defined by a UK 1990 birth weight reference standard(5). Perinatal death is defined as stillbirth or neonatal death. Perinatal morbidity is defined as ≥1 of the following: a 5 minute Apgar score <7, delivery with metabolic acidosis (defined as an umbilical cord artery or vein pH <7.1 and a base deficit of >10mmol/L), or admission to the neonatal unit (defined as admission <48 hours after birth and discharge ≥48 hours after admission). The second outcome is a subset of the first, namely, composite severe adverse outcome. This is defined as one or more of the following: (i) preeclampsia with severe features (ACOG 2013 definition), or (ii) perinatal death or severe perinatal morbidity. Severe perinatal morbidity is defined as livebirth associated with hypoxic ischaemic encephalopathy (any grade), use of inotropes, mechanical ventilation, or severe metabolic acidosis (defined as an umbilical cord artery or vein pH <7.0 and a base deficit of >12mmol/L). As described above, the definition of perinatal morbidity or mortality for both primary outcomes is itself a composite of different adverse perinatal outcomes, and the wording “the individual elements of the composite outcomes” refers to the latter set of outcomes.

6.1.2 Secondary outcomes

Secondary outcomes will be compared between the control and intervention arms of the women who screened high risk. As well as the individual elements of the composite primary outcomes, the full list of secondary outcomes includes a range of other outcomes which could plausibly be affected by intervention (Table 2).

**Table 2.** Secondary outcomes (maternal and offspring)

| **Maternal** | **Offspring** |
|---|---|
| Pre-eclampsia | Apgar at 5 minutes <7 |
| Pre-eclampsia with severe features | Cord blood metabolic acidosis (pH < 7.1 & base deficit >10mmol/L) |
| Death | Admission to SCBU or NICU for >48h duration and <48h following delivery |
| Stroke | Perinatal death |
| Admission to any specialist medical unit (ICU, CCU, NCCU) | Any perinatal morbidity or mortality (i.e. one or more of Apgar at 5 minutes <7, cord blood metabolic acidosis, admission to SCBU or NICU for >48h duration and <48h following delivery, or perinatal death) |
| Placental abruption | Birth weight <3rd percentile |
| Cord prolapse | Hypoxic ischaemic encephalopathy (any grade) |
| Caesarean section | Use of inotropes |
| Epidural anaesthesia | Use of mechanical ventilation |
| Instrumental vaginal delivery | Severe cord blood metabolic acidosis (pH < 7.0 & base deficit >12mmol/L) |
| General anaesthesia | Any severe perinatal morbidity or mortality (i.e. one or more of hypoxic ischaemic encephalopathy, use of inotropes, use of mechanical ventilation, severe cord blood metabolic acidosis or perinatal death) |
| Post-partum haemorrhage (any >500ml) | Apgar at 1 minute |

| | |
|---|---|
| Post-partum haemorrhage requiring blood transfusion | Apgar at 5 minutes |
| Infection (pyrexia >38$^{o}$C and antibiotics) | Umbilical arterial pH |
| | Umbilical arterial base deficit |
| | Umbilical vein pH |
| | Umbilical vein base deficit |
| | Any admission to Special Care |
| | Duration of admission to Special Care |
| | Any admission to Neonatal Intensive Care |
| | Duration of admission to Neonatal Intensive Care |
| | Fracture (any) |
| | Seizures |
| | Prolonged hypotonia |
| | Abnormal level of consciousness |
| | Tube feeding (any duration) |
| | Therapeutic hypothermia |
| | Phototherapy |
| | Severe hypoglycaemia (requiring parenteral treatment or drug therapy) |
| | Birth weight (g) |
| | Birth weight percentile for sex and gestational age |
| | Stage of hypoxic ischaemic encephalopathy |
| | Oxygen administration and/or respiratory support |

All maternal and perinatal outcomes relate to the delivery admission. i.e. maternal or perinatal outcomes associated with re-admission following the post-delivery discharge are not included in the above.

6.2 Analysis of efficacy of the screening test

6.2.1 Assessment of the diagnostic effectiveness of the screening test

Comparison of women in groups (b) and (c) (described in section 4.3 above) will allow assessment of the effectiveness of the screening test in relation to the two primary outcomes. All women who consent to the RCT will have the screening test performed. Women who screen positive will be randomly allocated to the intervention or control groups. The latter involves blinding the participant and the clinical care team to the result and the women receiving routine clinical care, as is the case for the low risk group. Hence, the effectiveness of the screening test can be assessed by comparing high-risk control women with the women who screened low risk. In these analyses, the high-risk controls will be weighted (using probability weights with robust standard errors) by the inverse of the sampling fraction from all women who screened high-risk (this will be two in the event that the two groups randomised are identical in size).

The primary analysis of the screening performance of the test is a binary assessment of risk status (i.e. high or low risk) in relation to the two primary outcomes, hence screening performance will be assessed by sensitivity, specificity, positive and negative predictive values, positive and negative likelihood ratios, and the diagnostic odds ratio. The screening performance of the test will also be analysed for the individual elements of the primary composite outcomes. In the analysis of perinatal morbidity and mortality, this analysis will be confined to the composite of different adverse perinatal outcomes. For example, the analysis of screening performance for the elements of the first primary outcome will consist of three analyses: (i) any diagnosis of preeclampsia, (ii) birth weight percentile <3$^{rd}$, and (iii) the perinatal composite defined as one or more of the following: perinatal death, a 5 minute Apgar score <7, an umbilical cord artery or vein pH <7.1 and a base deficit of >10mmol/L), or admission to the neonatal unit <48 hours after birth and discharge ≥48 hours after admission. Any further analysis of screening results will be labelled as hypothesis generating. When these include assessment of numerical predictors, discrimination will be assessed using the area under the receiver operating characteristic curve.

6.2.2 Handling missing data in the assessment of the diagnostic effectiveness of the screening test

The classification of being high risk is described in brief in section 6.2.3 below and in detail in the accompanying protocol paper. Women who are eligible for the RCT at the 36wkGA visit and give consent then receive the risk assessment. Women who screen high risk (predicted to be about 6% to 7% of those screened) are then randomised to intervention or control. At the time of consenting for the RCT, the clinician recruiting the participant checks that all of the maternal characteristics required for the risk assessment have been retrieved. Hence, we anticipate minimal or zero missing data in relation to the maternal characteristics required to classify women as high risk. In the event of missing data on maternal characteristics, these will be imputed as the given risk factor being absent. Risk status is also defined by ultrasonic information. As eligibility requires that the participant had the 36wkGA research ultrasound, we anticipate minimal or zero missing data in relation to the 36wkGA ultrasonic estimated fetal weight in defining risk status. In the event of missing estimated fetal weight data from the 36wkGA research ultrasound, this will be imputed as the baby having an estimated fetal weight at or above the 10th percentile. It is likely that the assessment of uterine artery Doppler will be impossible in a small proportion of patients. In these cases, an absent assessment of the normality of the 20wkGA assessment of uterine artery Doppler will be imputed as normal Doppler. There will be a small proportion of patients where it will not be possible to measure the sFLT1:PlGF ratio. These women are not eligible for randomisation and all records with an absent sFLT1:PlGF ratio will be dropped from the analysis of screening performance.

Where there is no information at all about the birth, comparison of screening efficacy will be by complete case analysis, i.e. women who were lost to follow up following the risk assessment will be dropped from the analysis. This analysis is valid under a missing at random assumption. If there is a >5% imbalance in missing outcome data between low risk women and high risk women randomised to control, we will also evaluate whether results are robust to multiple imputation under missing not at random assumptions.

Where outcome data were retrieved but a given diagnosis was not documented, either positively or negatively, the participant will be assumed to have not experienced the given complication. For example, it will not be possible to obtain umbilical cord blood gas analysis from all randomised women, hence in the records where no cord blood gas analysis is documented the neonate will be classified as not having the metabolic acidosis component of perinatal morbidity. The rate of missingness between low risk women and high risk women randomised to control will be assessed. If there is a >5% imbalance in the rate of missingness between the groups, missing values for the given element of the composite will also be imputed within each group (i.e. separately within the high risk control group and the low risk group) using multiple imputation.

6.2.3 Additional analyses in the assessment of the diagnostic effectiveness of the screening test

The high risk group will be classified into four strata based on the elements of the screening assessment. These are ranked in descending order and where a given participant falls into more than one of the categories they will be defined by the highest category. The strata are (in descending order): sFLT1:PlGF ratio>110, sFLT1:PlGF ratio >38 and <110 plus estimated fetal weight <10th percentile on ultrasound scan; sFLT1:PlGF ratio >38 and <110 plus high resistance uterine Doppler at ~20wkGA; and sFLT1:PlGF ratio >38 and <110 plus maternal risk factors. The positive predictive value of the screening test in the different strata of high-risk will be described by a forest plot of the proportion of women (and bars indicating 95% CI) who experienced the primary outcomes (separate plots for each of the primary outcomes) among the group who screened high-risk and were randomised to the control group (concealed result and routine care) in the different strata. The plot will also include the proportion of women experiencing the primary outcome in the women who screened low risk. The statistical significance of any variation across the four strata will be determined by a Chi squared test (or Fisher's exact test, if the expected number of observations in any cell is <5).

6.3 Analysis of efficacy of intervention

6.3.1 Assessment of the clinical effectiveness of the intervention

The RCT aims to study nulliparous women at around 36wkGA who are found to be high risk (using a novel combination of biochemical, ultrasonic and clinical assessment) of composite outcomes of preeclampsia, fetal growth restriction and neonatal morbidity and mortality, to determine whether the offer of earlier delivery and/or enhanced monitoring results in a risk ratio of less than one for the composite outcome, based on being randomised to this intervention referent to routine care, irrespective of the subsequent clinical management. Comparison of the rates of the composite outcomes in the high-risk group comparing women who were randomised to intervention with women randomised to routine care will be assessed by the adjusted risk ratio (RR) and 95% CI estimated using Poisson regression with robust standard errors, adjusted for stratification variables and for a range of other characteristics associated with the primary outcomes. All RRs will be for the group where the result was revealed referent to the group where the result was concealed.

6.3.2 Multivariate analysis of the assessment of the clinical effectiveness of the intervention

Stratification by the categories of the risk assessment approach will ensure similar compositions of both groups in terms of the reason for screening high-risk, however, the RR will be adjusted for the stratification group. Given the use of randomisation, imbalances in maternal characteristics are unlikely to be observed but could occur through the play of chance and have the potential to influence the apparent effect of intervention. Consequently, the statistical power of the analysis is likely to be enhanced by adjustment for characteristics which could affect the primary outcome. In order to select which characteristics to use in this adjustment, we fitted a Poisson model to the original POPS dataset with the composite primary outcome of POPS2 as the outcome, with the categorical stratification groups as covariates, and with the following characteristics added to these covariates (categorical, unless indicated otherwise):

- sFLT1:PlGF (log transformed ratio)
- Estimated fetal weight at 36 weeks [continuous, z score of the estimated fetal weight, adjusted for exact GA at measurement using the Hadlock 1991 reference(6)]
- 20wk uterine artery Doppler mean PI [continuous, z score of log-transformed ~20wk uterine artery Doppler mean PI, adjusted for exact GA at measurement using linear regression, as previously described(2)]
- Maternal age (continuous maternal age at recruitment)
- Maternal BMI (continuous, log-transformed BMI at visit 1)
- Chronic kidney disease
- Autoimmune disease such as systemic lupus erythematosus or antiphospholipid syndrome
- Type 1 or type 2 diabetes
- Chronic hypertension
- Age at discontinuing full time education (continuous)
- Ethnicity
- Self-reported smoking status (dichotomised: current smokers versus all others)
- Socio-economic deprivation status of postcode of residence
- Marital status

We included all of the covariates in the model. We then used backward stepwise regression (P threshold = 0.05) with the stratification groups forced into the model to identify which of the additional characteristics were significantly associated with the outcome in a model which included the stratification groups. This analysis identified the following characteristics as being associated with the primary outcome having already adjusted for the randomisation strata: (i) sFLT1:PlGF, (ii) estimated fetal weight at 36 weeks z score, based on the Hadlock 1991 analysis(6), (iii) chronic hypertension, (iv) self-reported smoking status, (v) maternal BMI. Hence, in the analysis of outcomes in POPS2 we will adjust for these variables in addition to the stratification groups.

6.3.3 Handling missing data in the assessment of the clinical effectiveness of the intervention
Missing outcome data where there is no information at all about the birth will be analysed by complete case analysis, i.e. women who were lost to follow up following randomisation will be dropped from the analysis. This analysis is valid under a missing at random assumption. For each primary and secondary outcome, if there is a >5% imbalance in missing outcome data between randomised groups, we will also evaluate whether results are robust to multiple imputation under missing not at random assumptions.

Where outcome data were retrieved but a given diagnosis was not documented, either positively or negatively, the participant will be assumed to have not experienced the given complication. For example, it will not be possible to obtain umbilical cord blood gas analysis from all randomised women, hence in the records where no cord blood gas analysis is documented the neonate will be classified as not having the metabolic acidosis component of perinatal morbidity. The rate of missingness between the two arms of the RCT for the missing elements of the composite will be assessed. If there is a ≤5% difference in the rate of missingness, missing values will be managed as described above, i.e. by assuming that the outcome was not observed. If there is a >5% imbalance in the rate of missingness between the groups, we will also multiply impute missing values for the given element of the composite using multiple imputation by randomised group (i.e. separately within the intervention and control groups), including in the imputation models all the other elements of the composites, the stratification groups, and the adjustment covariates (sFLT1:PlGF, estimated fetal weight at 36 weeks z score, chronic hypertension, self-reported smoking status, and maternal BMI). Missing baseline covariate data will be handled by mean imputation in the multivariate analysis.

6.3.4 Additional analyses in the assessment of the clinical effectiveness of the intervention

The adjusted risk ratio for the intervention will be calculated within each of the four strata (described in section 6.2.3) and plotted in a forest plot for the two primary outcomes. The statistical significance of apparent variation in the effect of the intervention across the four groups will be assessed by an interaction term with three degrees of freedom in the Poisson regression model.

6.3.5 Analysis of secondary outcomes in the assessment of the clinical effectiveness of the intervention

Analysis of secondary outcomes will be clearly separated from the analysis of primary outcomes. The same statistical methods will be employed in the analysis of binary secondary outcomes as described above for the primary outcome. Analyses will be adjusted for the stratification groups and for the factors which were shown to be associated with the primary outcome in the POPS dataset, as described above. For secondary outcomes that are numerical variables, statistical comparison will adjust for the same covariates as for the primary outcome and will be performed using multiple linear regression. Confidence intervals will be checked using bootstrapping where the variables are non-normally distributed.

6.4 Additional analyses outside scope of the SAP

POPS2 is a prospective cohort study with a nested RCT. We anticipate many additional analyses of the dataset, involving both analysis of the whole cohort and other nested studies, such as case cohort and case control studies. POPS had a very similar design and has resulted in analyses with these three different study designs (see, for example, Sovio et al 2015(2), Sovio et al 2020(7) and de Goffau et al 2019(8)). The nested RCT will, however, influence how these studies are designed. For example, in the case of analysing POPS2 as a cohort, any analysis will have to address the effect of the intervention in high-risk women. This could be accomplished by dropping women in group (d) from the analysis and weighting women in group (c) by two (i.e. the inverse of the sampling fraction assuming the number of women in the intervention and control groups are identical). However, description of these analyses lies outside the scope of the current document.

6.5 Harms

The definition and management of harm is described in accompanying protocol paper.

6.6 Statistical software

Stata (Statacorp, College Station, TX, USA) will be used for all statistical analyses, using version 19 or later.
