## Supplementary material for "A prospective cohort study of unselected nulliparous women with a nested randomized controlled trial of screening using the sFLT1:PlGF ratio, ultrasound and maternal characteristics and intervention using enhanced monitoring and early delivery: study protocol for the POPS2 cohort and randomized cont": SPIRIT checklist

| | | Reporting Item | Page and Line Number | Reason if not applicable |
|---|---|---|---|---|
| **Administrative information** | | | | |
| Title | #1 | Descriptive title identifying the study design, population, interventions, and, if applicable, trial acronym | Page 1, lines 1-4 | |
| Trial registration | #2a | Trial identifier and registry name. If not yet registered, name of intended registry | Page 2, line 55 | |
| Trial registration: data set | #2b | All items from the World Health Organization Trial Registration Data Set | Page 2, line 55 | |
| Protocol version | #3 | Date and version identifier | Page 1, line 5 | |
| Funding | #4 | Sources and types of financial, material, and other support | Page 31, lines 986-997 | |
| Roles and responsibilities: contributorship | #5a | Names, affiliations, and roles of protocol contributors | Page 32, lines 976-983 | |
| Roles and responsibilities: sponsor contact information | #5b | Name and contact information for the trial sponsor | Page 33, line 1054-1057 | |
| Roles and responsibilities: sponsor and funder | #5c | Role of study sponsor and funders, if any, in study design; collection, management, analysis, and interpretation of | Page 32, line 996-997 | |

| | | | |
|---|---|---|---|
| | | data; writing of the report; and the decision to submit the report for publication, including whether they will have ultimate authority over any of these activities | |
| Roles and responsibilities: committees | #5d | Composition, roles, and responsibilities of the coordinating centre, steering committee, endpoint adjudication committee, data management team, and other individuals or groups overseeing the trial, if applicable (see Item 21a for data monitoring committee) | Page 27, lines 815-847 |
| **Introduction** | | | |
| Background and rationale | #6a | Description of research question and justification for undertaking the trial, including summary of relevant studies (published and unpublished) examining benefits and harms for each intervention | Page 5, lines 62-104 |
| Background and rationale: choice of comparators | #6b | Explanation for choice of comparators | Page 5, lines 62-104 |
| Objectives | #7 | Specific objectives or hypotheses | Page 6, lines 106-121 |

| | | | |
|---|---|---|---|
| Trial design | #8 | Description of trial design including type of trial (eg, parallel group, crossover, factorial, single group), allocation ratio, and framework (eg, superiority, equivalence, non-inferiority, exploratory) | Page 6, lines 123-126 |
| **Methods: Participants, interventions, and outcomes** | | | |
| Study setting | #9 | Description of study settings (eg, community clinic, academic hospital) and list of countries where data will be collected. Reference to where list of study sites can be obtained | Page 7, lines 130-134 |
| Eligibility criteria | #10 | Inclusion and exclusion criteria for participants. If applicable, eligibility criteria for study centres and individuals who will perform the interventions (eg, surgeons, psychotherapists) | Page 7, lines 136-142 |
| Interventions: description | #11a | Interventions for each group with sufficient detail to allow replication, including how and when they will be administered | Page 11, lines 275-320 |
| Interventions: modifications | #11b | Criteria for discontinuing or modifying allocated interventions for a given | Page 12, lines 322-327 |

| | | | |
|---|---|---|---|
| | | trial participant (eg, drug dose change in response to harms, participant request, or improving / worsening disease) | |
| Interventions: adherance | #11c | Strategies to improve adherence to intervention protocols, and any procedures for monitoring adherence (eg, drug tablet return; laboratory tests) | Pages 12-13, lines 329-335 |
| Interventions: concomitant care | #11d | Relevant concomitant care and interventions that are permitted or prohibited during the trial | Page 13, lines 337-339 |
| Outcomes | #12 | Primary, secondary, and other outcomes, including the specific measurement variable (eg, systolic blood pressure), analysis metric (eg, change from baseline, final value, time to event), method of aggregation (eg, median, proportion), and time point for each outcome. Explanation of the clinical relevance of chosen efficacy and harm outcomes is strongly recommended | Page 13-14, lines 352-385 |
| Participant timeline | #13 | Time schedule of enrolment, interventions (including any run-ins and | Page 15, lines 387-510 |

| | | | |
|---|---|---|---|
| | | washouts), assessments, and visits for participants. A schematic diagram is highly recommended (see Figure) | |
| Sample size | #14 | Estimated number of participants needed to achieve study objectives and how it was determined, including clinical and statistical assumptions supporting any sample size calculations | Page 18, lines 512-566 |
| Recruitment | #15 | Strategies for achieving adequate participant enrolment to reach target sample size | Page 20, lines 568-580 |
| **Methods: Assignment of interventions (for controlled trials)** | | | |
| Allocation: sequence generation | #16a | Method of generating the allocation sequence (eg, computer-generated random numbers), and list of any factors for stratification. To reduce predictability of a random sequence, details of any planned restriction (eg, blocking) should be provided in a separate document that is unavailable to those who enrol participants or assign interventions | Page 20, lines 582-591 |

| | | | |
|---|---|---|---|
| Allocation concealment mechanism | #16b | Mechanism of implementing the allocation sequence (eg, central telephone; sequentially numbered, opaque, sealed envelopes), describing any steps to conceal the sequence until interventions are assigned | Pages 20-21, lines 598-607 |
| Allocation: implementation | #16c | Who will generate the allocation sequence, who will enrol participants, and who will assign participants to interventions | Page 21, lines 609-622 |
| Blinding (masking) | #17a | Who will be blinded after assignment to interventions (eg, trial participants, care providers, outcome assessors, data analysts), and how | Page 21, lines 624-633 |
| Blinding (masking): emergency unblinding | #17b | If blinded, circumstances under which unblinding is permissible, and procedure for revealing a participant's allocated intervention during the trial | Pages 21-22, lines 635-642 |
| **Methods: Data collection, management, and analysis** | | | |
| Data collection plan | #18a | Plans for assessment and collection of outcome, baseline, and other trial data, including any related | Page 22, lines 644-667 |

| | | | |
|---|---|---|---|
| | | processes to promote data quality (eg, duplicate measurements, training of assessors) and a description of study instruments (eg, questionnaires, laboratory tests) along with their reliability and validity, if known. Reference to where data collection forms can be found, if not in the protocol | |
| Data collection plan: retention | #18b | Plans to promote participant retention and complete follow-up, including list of any outcome data to be collected for participants who discontinue or deviate from intervention protocols | Pages 22-23, lines 669-681 |
| Data management | #19 | Plans for data entry, coding, security, and storage, including any related processes to promote data quality (eg, double data entry; range checks for data values). Reference to where details of data management procedures can be found, if not in the protocol | Page 23, lines 683-697 |

| | | | |
|---|---|---|---|
| Statistics: outcomes | #20a | Statistical methods for analysing primary and secondary outcomes. Reference to where other details of the statistical analysis plan can be found, if not in the protocol | Page 25, lines 759-774 |
| Statistics: additional analyses | #20b | Methods for any additional analyses (eg, subgroup and adjusted analyses) | Page 25, lines 759-774 |
| Statistics: analysis population and missing data | #20c | Definition of analysis population relating to protocol non-adherence (eg, as randomised analysis), and any statistical methods to handle missing data (eg, multiple imputation) | Page 25, lines 759-774 |
| **Methods: Monitoring** | | | |
| Data monitoring: formal committee | #21a | Composition of data monitoring committee (DMC); summary of its role and reporting structure; statement of whether it is independent from the sponsor and competing interests; and reference to where further details about its charter can be found, if not in the protocol. Alternatively, an explanation of why a DMC is not needed | Pages 27-28, lines 837-847 |

| | | | |
|---|---|---|---|
| Data monitoring: interim analysis | #21b | Description of any interim analyses and stopping guidelines, including who will have access to these interim results and make the final decision to terminate the trial | Page 26, lines 776-788 |
| Harms | #22 | Plans for collecting, assessing, reporting, and managing solicited and spontaneously reported adverse events and other unintended effects of trial interventions or trial conduct | Pages 28-29, lines 849-874 |
| Auditing | #23 | Frequency and procedures for auditing trial conduct, if any, and whether the process will be independent from investigators and the sponsor | Page 29, lines 876-881 |
| **Ethics and dissemination** | | | |
| Research ethics approval | #24 | Plans for seeking research ethics committee / institutional review board (REC / IRB) approval | Page 32, lines 1006-1012 |
| Protocol amendments | #25 | Plans for communicating important protocol modifications (eg, changes to eligibility criteria, outcomes, analyses) to relevant | Page 29, lines 883-893 |

| | | | |
|---|---|---|---|
| | | parties (eg, investigators, REC / IRBs, trial participants, trial registries, journals, regulators) | |
| Consent or assent | #26a | Who will obtain informed consent or assent from potential trial participants or authorised surrogates, and how (see Item 32) | Pages 7-10 lines 153-259 |
| Consent or assent: ancillary studies | #26b | Additional consent provisions for collection and use of participant data and biological specimens in ancillary studies, if applicable | Pages 10-11, lines 261-273 |
| Confidentiality | #27 | How personal information about potential and enrolled participants will be collected, shared, and maintained in order to protect confidentiality before, during, and after the trial | Pages 23-24, lines 699-726 |
| Declaration of interests | #28 | Financial and other competing interests for principal investigators for the overall trial and each study site | Page 33, lines 1017-1035 |
| Data access | #29 | Statement of who will have access to the final trial dataset, and disclosure of contractual agreements that limit such access for investigators | Page 27, lines 808-813 |

| | | | |
|---|---|---|---|
| Ancillary and post trial care | #30 | Provisions, if any, for ancillary and post-trial care, and for compensation to those who suffer harm from trial participation | Page 13, lines 341-350 |
| Dissemination policy: trial results | #31a | Plans for investigators and sponsor to communicate trial results to participants, healthcare professionals, the public, and other relevant groups (eg, via publication, reporting in results databases, or other data sharing arrangements), including any publication restrictions | Page 29, lines 895-903 |
| Dissemination policy: authorship | #31b | Authorship eligibility guidelines and any intended use of professional writers | Page 29, lines 895-903 |
| Dissemination policy: reproducible research | #31c | Plans, if any, for granting public access to the full protocol, participant-level dataset, and statistical code | Page 27, lines 808-813 |
| **Appendices** | | | |
| Informed consent materials | #32 | Model consent form and other related documentation given to participants and authorised surrogates | Page 33, lines 1014-1015 |

| | | | |
|---|---|---|---|
| Biological specimens | #33 | Plans for collection, laboratory evaluation, and storage of biological specimens for genetic or molecular analysis in the current trial and for future use in ancillary studies, if applicable | Page 24, lines 728-757 |

It is strongly recommended that this checklist be read in conjunction with the SPIRIT 2013 Explanation & Elaboration for important clarification on the items. Amendments to the protocol should be tracked and dated. The SPIRIT checklist is copyrighted by the SPIRIT Group under the Creative Commons "Attribution-NonCommercial-NoDerivs 3.0 Unported" license. This checklist can be completed online using https://www.goodreports.org/, a tool made by the EQUATOR Network in collaboration with Penelope.ai
